# Drug-resistant focal epilepsy in children is associated with increased modal controllability of the whole brain and epileptogenic regions

**DOI:** 10.1101/2021.08.05.21261484

**Authors:** Aswin Chari, Kiran Seunarine, Xiaosong He, Martin Tisdall, Christopher A Clark, Danielle S Bassett, Rodney C Scott, Richard E Rosch

## Abstract

Network control theory provides a framework by which neurophysiological dynamics of the brain can be modelled as a function of the structural connectome constructed from diffusion MRI. Average controllability describes the ability of a region to drive the brain to easy-to-reach neurophysiological states whilst modal controllability describes the ability of a region to drive the brain to difficult-to-reach states.

In this study, we identify increases in mean average and modal controllability in children with drug-resistant epilepsy compared to healthy controls. Using simulations, we purport that these changes may be a result of increased thalamocortical connectivity. At the node level, we demonstrate decreased modal controllability in the thalamus and posterior cingulate regions. In those undergoing resective surgery, we also demonstrate increased modal controllability of the resected parcels, a finding specific to patients who were rendered seizure free following surgery.

Changes in controllability are a manifestation of brain network dysfunction in epilepsy and may be a useful construct to understand the pathophysiology of this archetypical network disease. Understanding the mechanisms underlying these controllability changes may also facilitate the design of network-focussed interventions that seek to normalise network structure and function.

## Introduction

Disruptions to structural and functional brain networks are systems-level mechanisms of many neurological and psychiatric disorders. Structural networks are commonly assessed using diffusion-weighted magnetic resonance imaging (DWI) whereas functional networks can be assessed with functional magnetic resonance imaging (fMRI), electroencephalography (EEG), magnetoencephalography (MEG) or intracranial electroencephalography (iEEG) techniques, among others. Epilepsy is one of the common neurological disorders associated with measurable network dysfunction. A variety of different modalities have been used to examine mechanisms of seizure dynamics and associated comorbidities, investigate the effects of treatment and, increasingly, to guide the development of optimal management strategies. ^1–12^ Surgical treatments have been shown to alter whole-brain network dynamics, but the implications of these changes for ongoing seizure risk and cognitive outcomes are yet to be fully elucidated.^10,13^

In children with epilepsy, many of whom have concurrent cognitive and psychological comorbidities, the acquisition of functional network data, particularly in the awake state, may be difficult. In light of recent advances in systems engineering, the use of structural brain networks to model neurophysiological dynamics is an attractive alternative.^4,11,12^ More specifically, the concept of network control theory has been applied to the study of brain networks, describing how the structure of a networked system such as the brain, both supports and constrains functional dynamics.^14,15^ It provides an understanding of the networked system’s response to internal or external perturbation.^14–16^ As applied to the brain, control theory metrics can be calculated from structural connectivity matrices derived from *in-vivo* diffusion MRI, after assuming a particular form for the network’s dynamics.^15^ The nodes are anatomically defined parcels of grey matter and the weight of the (i,j)-th edge in this undirected network is the number of white matter streamlines estimated between the parcel i and parcel j.

For the purposes of the control theory framework, we consider a ‘brain state’ to be a pattern of parcel activity; a single state is the magnitude of inferred neurophysiological activity across brain regions at a single time point. The temporal evolution of brain states can be modelled as a process of dynamics on the structural network. Aspects of this relationship between brain structure and dynamics can be captured by two controllability metrics: **average controllability** and **modal controllability** (Figure 1).^14,15^ Average controllability is used to measure the ability of a region to drive the brain to easy-to-reach states; in humans, regions of high average controllability tend to be densely connected, are involved in maintaining smooth and frequent transitions between related brain states and are enriched in the default mode network. Modal controllability is defined as the ability of a brain region to drive the system to a difficult-to-reach state; in humans, regions of high modal controllability tend to be weakly connected areas, allow switching between brain states that have high energy barriers between them, and are enriched in frontoparietal and cingulo-opercular control systems.^14,15^ In brain networks, an inverse relationship between these two measures of regional controllability has been reported, that is not necessarily present in other network types.^15,17^ This fact suggests parcels which can heavily modulate easy-to-reach states have a limited impact on the modulation of difficult-to-reach states and vice-versa. It is likely that this balance plays an important role in supporting and constraining brain dynamics.

**Figure 1:**
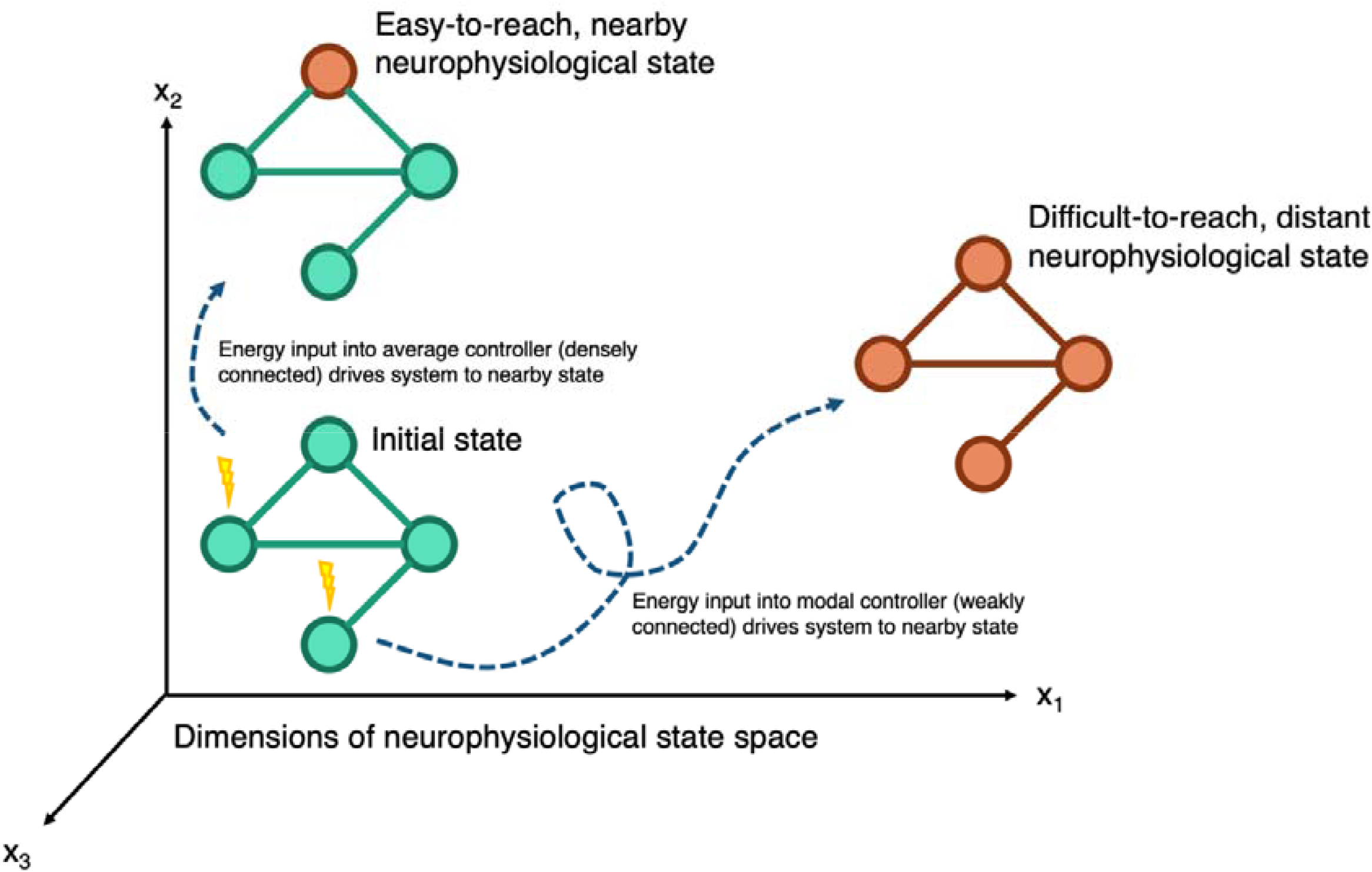
Schematic of network control theory, illustrating concepts of average and modal controllability. Brain networks exist in a neurophysiological state space, where the state is defined as the inferred magnitude of neurophysiological activity at a given time. Nodes with high average controllability tend to be densely connected and drive the system to nearby, easy-to-reach neurophysiological states. Nodes with high modal controllability tend to be weakly connected and drive the system to distant, difficult-to-reach neurophysiological states.

Disruption of these structural constraints of brain dynamics could be mechanisms driving outcomes in people with epilepsy.^18,19^ Existing reports of controllability in epilepsy are scarce. Bernhardt *et al*. demonstrated lower average controllability in the affected hemispheres of patients with surgically treated temporal lobe epilepsy, potentially accounting for the cognitive deficits seen in these patients. ^20^ In a complementary study, Scheid *et al*. used a time-evolving controllability as a framework to potentially guide the optimal location and minimal energy inputs for neuromodulation treatments.^21^ Here, we sought to understand changes in controllability in a cohort of children with surgically treated drug-resistant epilepsy at a single centre. Given the propensity of the epileptic brain to enter difficult-to-reach states of pathological neurophysiological activity that manifest as seizures, we hypothesized that children with focal drug-resistant epilepsy would have a higher modal controllability than healthy controls and that those with multifocal epilepsies would have even higher modal controllability than those with unifocal disease. Our second aim was to develop an understanding of how changes in controllability arise at the network structure level. We investigated brain network organisation in patients and healthy controls by studying the relationship between average and modal controllability, hypothesizing that there would be a weakening of this relationship resulting in a disorganisation of the highly specialised brain network architecture in patients with drug resistant epilepsy. Importantly, we sought to understand whether these changes were explained by changes in established graph metrics and sought to recreate these changes in the structural connectivity matrices through simulations. Our final aim was to study changes in controllability at the regional level in individual parcels. For patients undergoing resective surgery, we hypothesized that the modal controllability of the putative epileptogenic zone that underwent resection may be higher than corresponding areas in the controls. In testing and validating these hypotheses, our study adds weight to the concept of network-focussed interventions in drug-resistant epilepsy and, specifically the ability to study functional dynamics from readily available structural data.

## Results

### Demographics

A total of 16 healthy controls, 52 patients undergoing resective surgery for presumed unifocal epilepsy, and 27 patients undergoing insertion of vagal nerve stimulator (VNS) for presumed multifocal epilepsy were included (age range 3-20, see Supplementary Table 1). All patients had drug-resistant epilepsy^22^ and had been discussed at the epilepsy multidisciplinary team meeting at Great Ormond Street Hospital for Children. Subjects in the resective surgery group had a mixture of temporal and extratemporal resections and were classed as seizure free (Engel Class I, 33/52) or not seizure free (Engel Classes II-IV, 19/52) at last follow-up (median 1.5 years, range 0.6-4.1 years). Subjects in the VNS group were classed as responders (≥50% reduction in seizures, 11/27) and non-responders (<50% reduction in seizures, 16/27) at last follow-up (median 1.9 years, range 0.9-3.0 years). The distributions of age (Figure 2a, Kruskal-Wallis Test, p<0.0001) and baseline cognitive function measured by the pre-operative full-scale IQ (Figure 2b, available in 86/95 subjects, Chi-Sq Test, p=0.01) were not matched between the groups and were accordingly used as a covariate in subsequent analyses.

**Figure 2:**
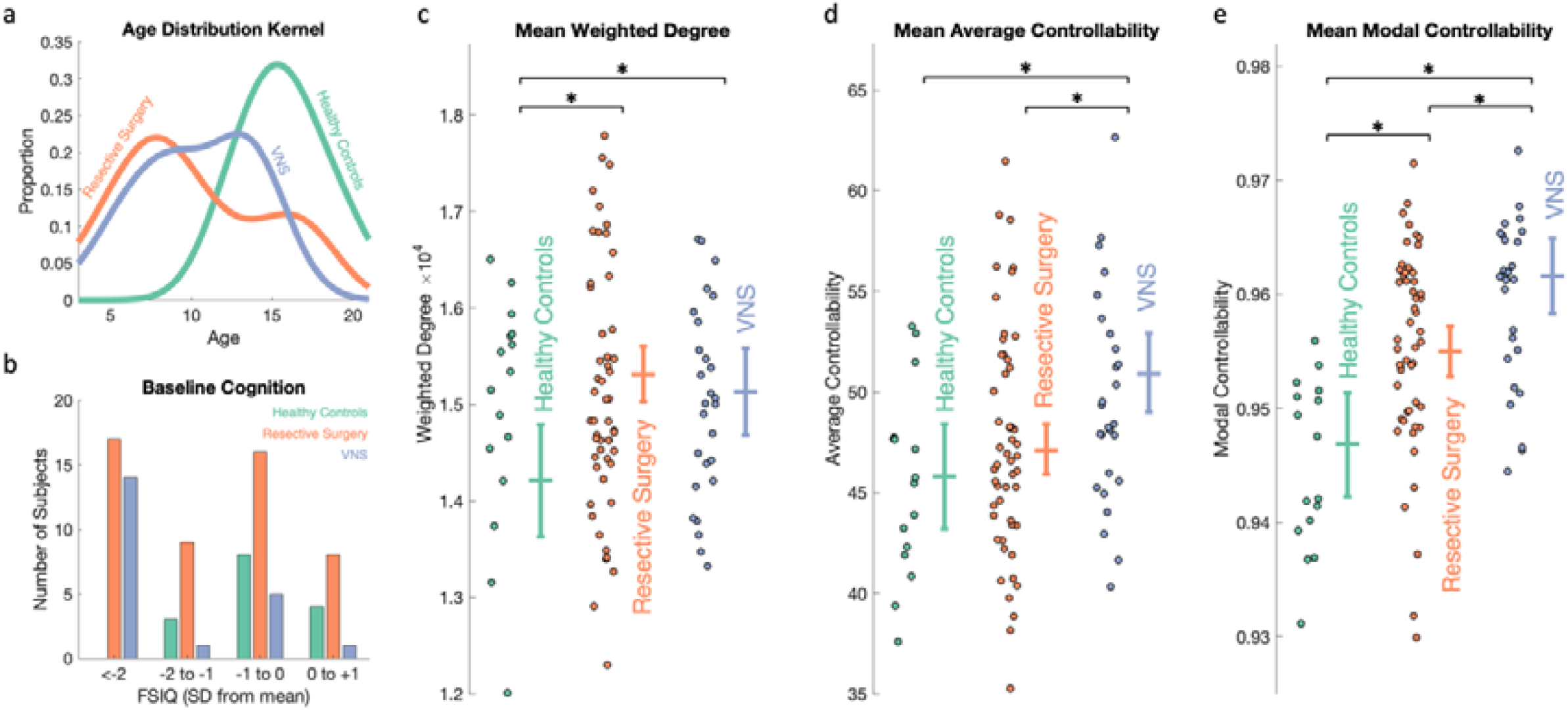
Drug-resistant epilepsy is associated with higher mean weighted degree, average controllability and modal controllability compared to healthy controls. (a) Kernel plot of age distributions which were significantly different between the groups (Kruskal-Wallis Test, p<0.0001). (b) Baseline cognitive function measured by full scale IQ (FSIQ) classes compared to age-corrected normal population. More epilepsy patients (resective surgery and VNS) were in the lower classes (Chi-Sq Test, p=0.01). (c-e) Scatter plots of (c) mean weighted degree, (d) mean average controllability, and (e) mean modal controllability for each group. Beside the scatter plots are estimated marginal means (+/− 95% Wald confidence intervals) following correction for age, sex, cognitive function and, for the controllability metrics, mean weighted degree. Multiple comparisons were adjusted for by least significant difference. Both groups of epilepsy patients had a higher weighted degree compared to controls (p=0.001 for resective surgery; p=0.02 for VNS groups). The VNS group had significantly higher mean average controllability compared to both the controls (p=0.003) and the resective surgery (p=0.001) group. Both groups had higher mean modal controllability compared to controls (p=0.003 for resective surgery; p=1×10^−6^ for VNS) and the VNS group was even significantly higher than the resective surgery group (p=0.001).

### Patients with epilepsy have higher mean weighted degree, average controllability, and modal controllability than healthy controls

To test whether there were group differences in network controllability at the whole brain level, we constructed structural connectivity matrices from pre-operative imaging derived from the number of streamlines connecting each of 253 cortical and subcortical parcels for each subject (see Methods for details). Weighted degree (defined as the sum of weights of edges connected to the node), average controllability, and modal controllability averaged across all parcels for each subject were compared between the groups using a generalised linear model (GLM) approach, correcting for age, sex, cognition and, for the controllability metrics, mean weighted degree and adjusted for multiple comparisons using Fisher’s least significant difference method (see Methods for details). Both groups of patients with epilepsy had a higher mean weighted degree compared to controls (p=0.001 for resective surgery; p=0.02 for VNS)(Figure 2c). The VNS group had a higher mean average controllability compared to controls (p=0.003) and resective surgery patients (p=0.001)(Figure 2d). Both groups had higher mean modal controllability compared to controls (p=0.003 for resective surgery; p=1×10^−6^ for VNS) and the VNS group had a higher mean modal controllability than the resective surgery group (p=0.001)(Figure 2e). These findings confirmed our hypothesis that children with drug-resistant epilepsy had higher modal controllability compared to healthy controls, indicating an increased propensity to reach distant states such as those associated with seizures. In agreement with previous studies, this whole brain increase in modal controllability was associated with higher whole-brain average controllability.^23^

### The node-level relationships between weighted degree, average controllability and modal controllability are less strong in children with drug-resistant epilepsy

To understand the regional variation underlying the differences in average and modal controllability, we examined the relationships between weighted degree, average controllability, and modal controllability at the level of the individual parcel. Previous studies have demonstrated that although the relationship between average and modal controllability can be negative, positive or non-significant at whole-network level, at the node-level there is a direct relationship between weighted degree and average controllability, and an inverse relationship between weighted degree and modal controllability.^15^ We hypothesized that patients with epilepsy would have weaker relationships between weighted degree, average controllability and modal controllability, indicating a loss of the network architecture that permits the specialisation of hubs for either average or modal control.^17^

To investigate these relationships, we converted the heavy-tailed raw weighted degree and controllability values to ranks in each patient (Supplementary Figure S1). Next, we calculated correlation coefficients to assess the strength of the relationship between the ranks of weighted degree and average controllability (WD-AC correlation), ranks of weighted degree and modal controllability (WD-MC correlation), and ranks of average and modal controllability (AC-MC correlation). Correlation coefficients were calculated for each subject and compared between groups correcting for age, sex, cognitive function and mean weighted degree. The WD-AC correlation coefficients were not different between the two groups of patients and controls (p=0.31 for resective surgery and p=0.10 for VNS). There was a significant difference in WD-MC correlation coefficients between the two groups and controls (p=0.02 for resective surgery and p=0.001 for VNS). Both groups also had a significantly weaker AC-MC correlation compared to the healthy controls (p = 7×10^−8^ for resective surgery and p=0.02 for VNS; see Figure 3). Together, these results indicate an alteration of the structural brain networks in patients with epilepsy that affects the separation of hubs with high average controllability and those with high modal controllability into non-overlapping brain areas.

**Figure 3:**
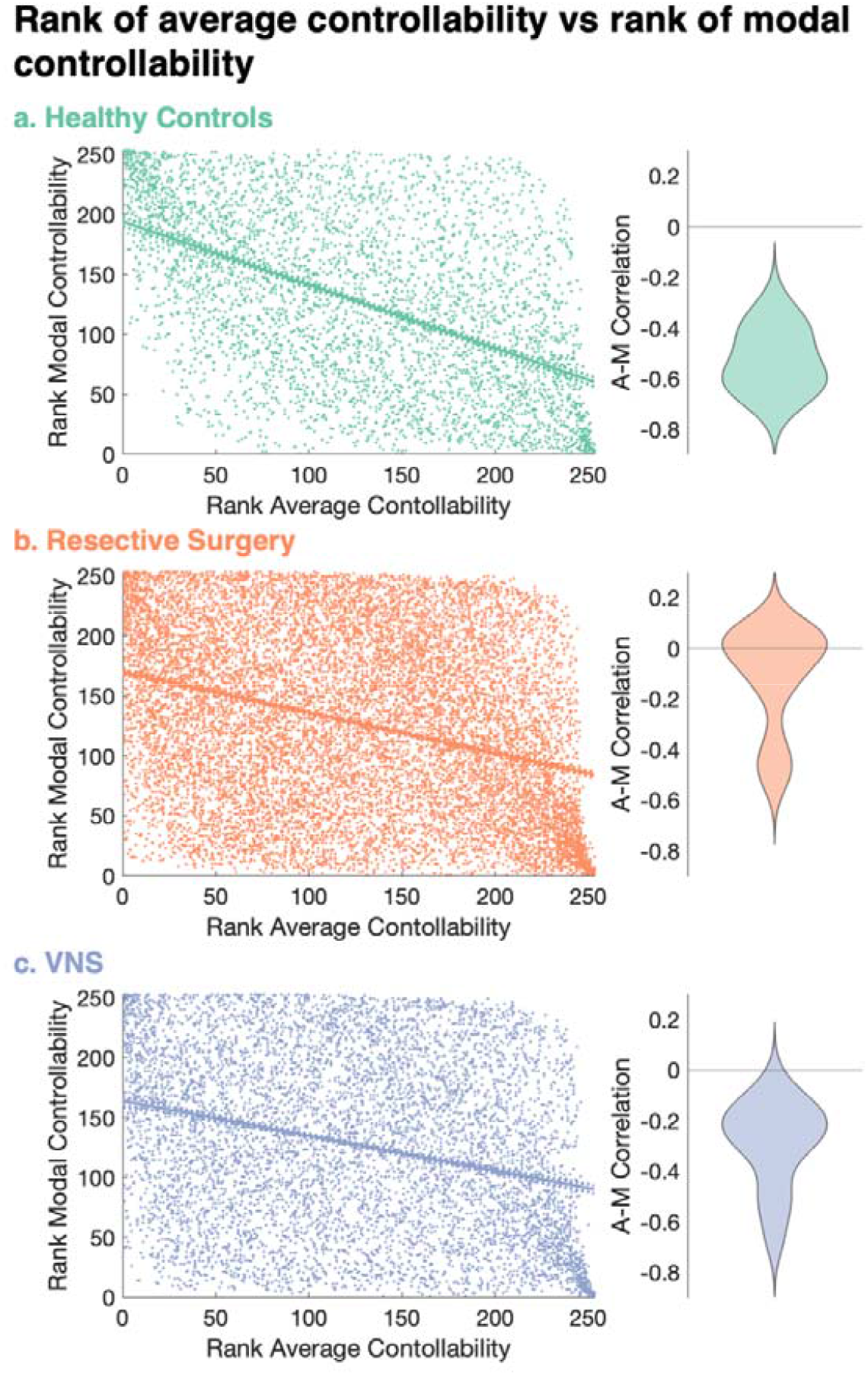
The inverse correlation between average controllability and modal controllability is weaker in patients with drug-resistant epilepsy. (a-c) Correlation between the rank of average controllability and rank of modal controllability (AC-MC correlation) for each parcel across all (a) healthy controls, (b) resective surgery, and (c) VNS patients. Lines show the linear regression best fit (solid line) and 95% confidence intervals (dotted lines). Adjacent violin plots show distribution of these correlation coefficients for each individual subject. There were significant differences in the distributions of the correlations at the individual patient level between controls and both groups of patients (p=7×10^−8^ for resective surgery and p=0.02 for VNS after correcting for age, sex, cognitive function, mean weighted degree and multiple comparisons).

### The controllability changes are not explained by graph measures but are explained by additional thalamocortical connections in the epileptic brain

In order to assess whether the network-level changes in controllability were merely reflective of changes in existing graph metrics that have been associated with drug-resistant epilepsy, we compared three measures, namely modularity, global efficiency and diffusion efficiency (see Methods for definitions) between groups.^24^ Compared to healthy controls, both groups of patients had a lower modularity (p=9×10^−6^ for resective surgery and p=4×10^−6^ for VNS) and higher diffusion efficiency (p=5×10^−7^ for both groups), but no difference in global efficiency (all metrics adjusted for age, sex, cognitive function and mean weighted degree and p-values were corrected for multiple comparisons). To gather a conceptual appreciation of the relationship between controllability metrics and the graph metrics, we plotted the correlation between the metrics (Supplementary Figure S2). On visual inspection, the groups segregated most clearly by the AC-MC correlation, and this segregation did not track strongly with any of the graph metrics.

One of the keys to further our understanding of network changes in epilepsy is to dissect how a healthy network can be altered into an epileptogenic one such that we can design novel therapeutics at any level of the complex systems framework to reverse these changes. We therefore sought to probe what the changes were in the structural connectivity matrices that resulted in these differences in controllability and node-level correlations between patients and controls. We started by assessing the density of the structural connectivity matrices (see Methods for definition). The healthy controls had an estimated marginal mean density of 0.63, compared to 0.71 in the resective surgery group (p = 3×10^−6^) and 0.69 in the VNS group (p=0.001) after correcting for age, sex, cognitive function and mean weighted degree and multiple comparisons. Kernel density distributions of edge weights showed minimal differences between the groups (Supplementary Figure S3). Together with the increase in weighted degree identified previously, these findings suggested an increased number of edges in the structural networks of children with epilepsy compared to healthy controls.

We therefore used numerical simulations to test whether the addition of edges at random to the healthy control connectivity matrix could recreate the specific network-wide alterations observed in our patient groups. To assess the impact on the network, we used the graph and controllability metrics as a ‘network fingerprint’. Adding edges at random led to an increase in average controllability but no significant change in modal controllability or the WD-AC, WD-MC and AC-MC correlation coefficients, inconsistent with our biological findings (Supplementary Figure S4).

We therefore hypothesized that the ‘network fingerprint’ observed in our data may be due to more anatomically constrained addition of edges. To investigate this further, we assessed the differences in mean connectivity matrices between controls and both groups of patients, identifying an increase in low streamline count (<100) edges, a majority of which were localised to ipsilateral thalamocortical connections (Supplementary Figure S5). To assess whether increases in thalamocortical connections alone were sufficient to recreate the changes in ‘network fingerprint’ changes observed in our patient cohort, we conducted a further simulation by increasing a fixed number (~4.5% of the total edges) of these ipsilateral thalamocortical connection edge weights in control subjects. We compared them to the baseline connectomes and null models of increasing an equal number of edges by equal weight anywhere in the ipsilateral hemisphere (see Methods for details). The anatomically constrained models reproduced the ‘network fingerprint’ changes seen in people with epilepsy more closely than the unconstrained null models (Figure 4). Specifically, compared to the controls, the anatomically constrained thalamocortical models had statistically significant increases in mean average controllability (p<1×10^−10^), mean modal controllability (p = 3×10^−9^), WD-MC correlation (p<1×10^−10^), AC-MC correlation (p=5×10^−4^), global efficiency (p=9×10^−9^), diffusion efficiency (p<1×10^−10^) and a decrease in modularity (p<1×10^−10^) after correcting for multiple comparisons (Benjamini-Hochberg FDR method). Interestingly, although the statistically significant findings were similar between the controls and the unconstrained null models, there was no significant difference in the AC-MC correlation (p=0.85 after correcting for multiple comparisons). This observation shows that the changes, specifically the increase in AC-MC correlation coefficient, a weakening of the negative correlation, observed in our patient data were most closely recreated by the anatomically constrained models that selectively increased thalamocortical weights.

**Figure 4:**
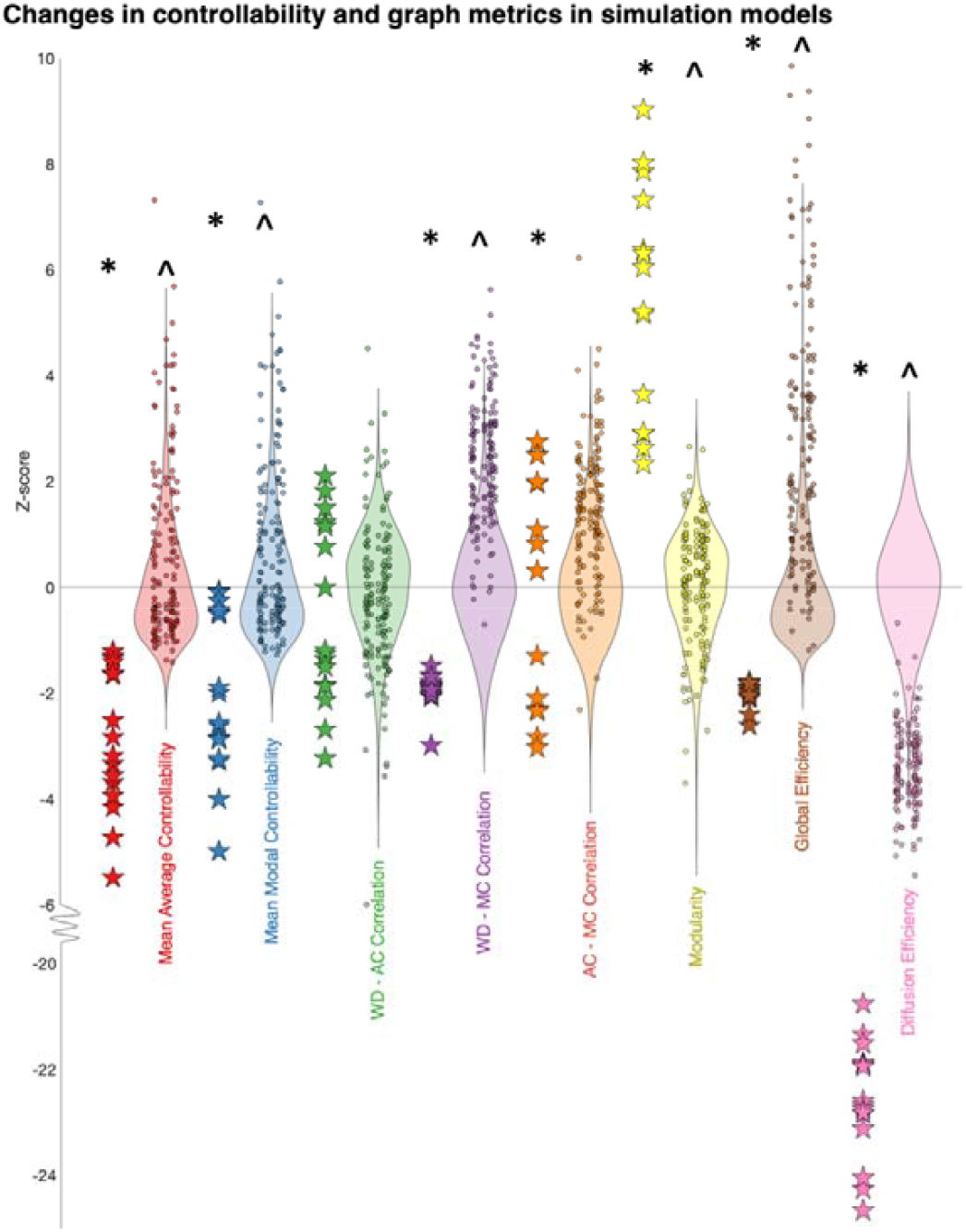
Changes in controllability and graph metrics observed in patient data is closely recreated by selectively increasing the weight of thalamocortical connections by a low number of streamlines. Z-scored controllability and graph metrics are compared between controls (stars) and anatomically-constrained simulations (dots, 10 for each control) of increasing ipsilateral thalamocortical edges (increase in edge weight of 4.5% (1440/31818) of edges by a streamline chosen from a normal distribution with *μ* = 50, *σ* = 15). Thebackground violin plots show the null models of increase in the same number of edges by the same weight, constrained to all ipsilateral edges only (200 for each control), on which the Z-scores were calculated. The distributions between the controls and constrained thalamocortical models (*) and controls and unconstrained null models (^) were compared using 2 sample t-tests and adjusted for multiple comparisons (Benjamini-Hochberg FDR method). Compared to the controls, the anatomically constrained thalamocortical models had statistically significant increases in mean average controllability (p<1×10^−10^), mean modal controllability (p=3×10^−9^), WD-MC correlation (p<1×10^−10^), AC-MC correlation (p=5×10^−4^), global efficiency (p=9×10^−9^), diffusion efficiency (p<1×10^−10^) and a decrease in modularity (p<1×10^−10^). Compared to the controls, the anatomically unconstrained models had statistically significant increases in mean average controllability (p<1×10^−10^), mean modal controllability (p<1×1-10^−10^), WD-MC correlation (p<1×10^−10^), global efficiency (p<1×10^−10^), diffusion efficiency (p<1×10^−10^) and a decrease in modularity (p<1×10^−10^). The specific important differentiator in our patient group (AC-MC correlation) was selectively replicated only in the anatomically constrained thalamocortical model.

### Global controllability metrics do not strongly associate with treatment outcome

Next, we sought to assess whether the controllability measures were markers for treatment success, both in the VNS and resective surgery groups (Supplementary Figure S6). After correction for age, sex, cognitive function, mean weighted degree and multiple comparisons, we observed subtle differences in the metrics, with the VNS non-responders having a higher mean average controllability than the responders (p=0.02) and the seizure-free patients following surgical resection having a higher mean modal controllability compared to the non-seizure-free patients (p<0.05). The weighted degree and correlation metrics were not discriminatory between groups once stratified by outcome.

### Thalamic & posterior cingulate parcels show increased weighted degree & decreased modal controllability in patients with drug-resistant epilepsy

In order to assess whether there were common nodes of the brain (e.g., thalamic nuclei as informed by the simulations) that subserved the changes in mean average and modal controllability, we created a Z-score for the ranks of each parcel in each patient using the mean and standard deviation from the controls; this procedure normalised the data (Supplementary Figure S1). Using a Z-score threshold of +/− 3.1 that served to correct for the 253 individual comparisons, there was significantly higher weighted degree and lower modal controllability of select bilateral thalamic nuclei and posterior/isthmus cingulate cortices in both groups of patients compared to controls (Table 1, Figure 5). This corroborated the findings of the simulations of increased thalamocortical connectivity that manifest in the control theory framework as lower modal controllability.

**Figure 5:**
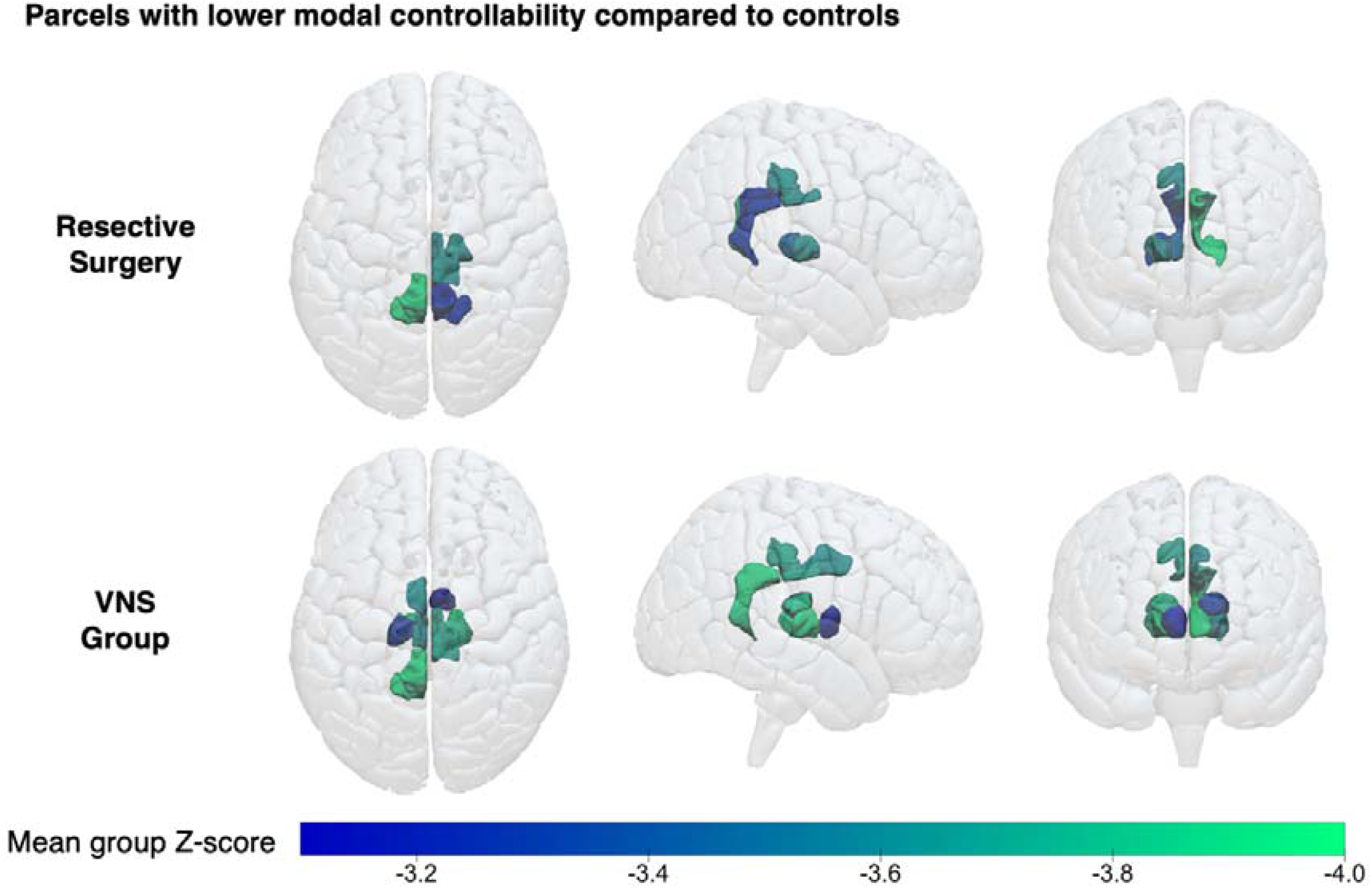
Patients with drug-resistant epilepsy show lower modal controllability of thalamic and posterior cingulate parcels. Visual maps of the parcels with significantly lower modal controllability (mean Z-scores <-3.1) in the resective surgery and VNS groups. Note the concentration of parcels around the thalamic nuclei and posterior cingulate cortices. The exact parcels are detailed in Table 1.

**Table 1:** Identified parcels that had a significantly different weighted degree, average controllability and modal controllability in both groups of patients compared to healthy controls. Parcels that had a significantly different (average Z-score threshold of +/− 3.1) weighted degree, average controllability and modal controllability for the resective surgery and VNS cohorts.

|  | Weighted Degree > Controls | Weighted Degree < Controls | Average Controllability > Controls | Average Controllability < Controls | Modal Controllability > Controls | Modal Controllability < Controls |
| --- | --- | --- | --- | --- | --- | --- |
| **Resective Surgery** | - Right Pulvinar<br>- Right Central-Lateral, Lateral-Posterior, Medial-Pulvinar<br>- Left Isthmus Cingulate 1<br>- Left-Pulvinar<br>- Left Central-Lateral, Lateral-Posterior, Medial-Pulvinar |  |  |  |  | - Right Posterior Cingulate 2<br>- Right Isthmus Cingulate 1<br>- Right Central-Lateral, Lateral-Posterior, Medial-Pulvinar<br>- Right Ventral-Latero-Ventral<br>- Left Isthmus Cingulate 1 |
| **VNS** | - Right-Pulvinar<br>- Right Ventral-Latero-Dorsal<br>- Right Central-Lateral, Lateral-Posterior, Medial-Pulvinar<br>- Left Posterior Cingulate<br>- Left Isthmus Cingulate 1<br>- Left Pulvinar<br>- Left Central-Lateral, Lateral-Posterior, Medial-Pulvinar | - Left Pericalcarine 1 | - Left Lateral Orbitofrontal 2<br>- Left Hypothalamus | - Right Lateral Occipital |  | - Right Posterior Cingulate 2<br>- Right Anterior Thalamus<br>- Right Ventral-Latero-Dorsal<br>- Right Central-Lateral, Lateral-Posterior, Medial-Pulvinar<br>- Right Ventral-Latero-Ventral<br>- Left Posterior Cingulate 1<br>- Left Isthmus Cingulate 1<br>- Left Medio-Dorsal<br>- Left Ventral-Latero-Dorsal<br>- Left Ventral- Latero-Ventral |

### Resected parcels show decreased weighted degree & increased modal controllability in patients undergoing resective surgery

Given the decreased thalamic and posterior cingulate modal controllability at the group level, we postulated that cortical parcels could have an increased modal controllability. We specifically hypothesized that this could be localised to parcels that are part of the putative epileptogenic zone that undergo resection. To test this, we identified the resected parcels from the post-operative scans and compared the mean Z-scores for the resected and non-resected parcels (see Methods for details, Supplementary Figure S7). The Z-scores were calculated using the mean and standard deviations of the ranks from the healthy controls; for the set of parcels in the control group, the mean Z-score would therefore be 0. In the resective surgery group, following correction for multiple comparisons, we found significant differences in the mean weighted degree and mean modal controllability between the whole brain and resected/non-resected parcels, only in patients that were seizure free following surgery (Figure 6a). Seizure free patients had a lower mean weighted degree (p=0.002) and higher mean modal controllability (p=6×10^−4^) of the resected parcels compared to the whole brain. A similar effect was observed when comparing the actual resections against 1000 ‘virtual resection’ null models of other randomly selected cortical parcels (Figure 6d, p=0.002 for weighted degree and p=0.007 for modal controllability), indicating that this effect was specific to the clinically identified putative epileptogenic zone. There were no significant differences between the means of the resected parcels in different histological groups (Kruskal Wallis tests, p=0.14 for weighted degree, p=0.15 foraver age controllability and p=0.56 for modal controllability; groups detailed in Supplementary Table 1).

**Figure 6:**
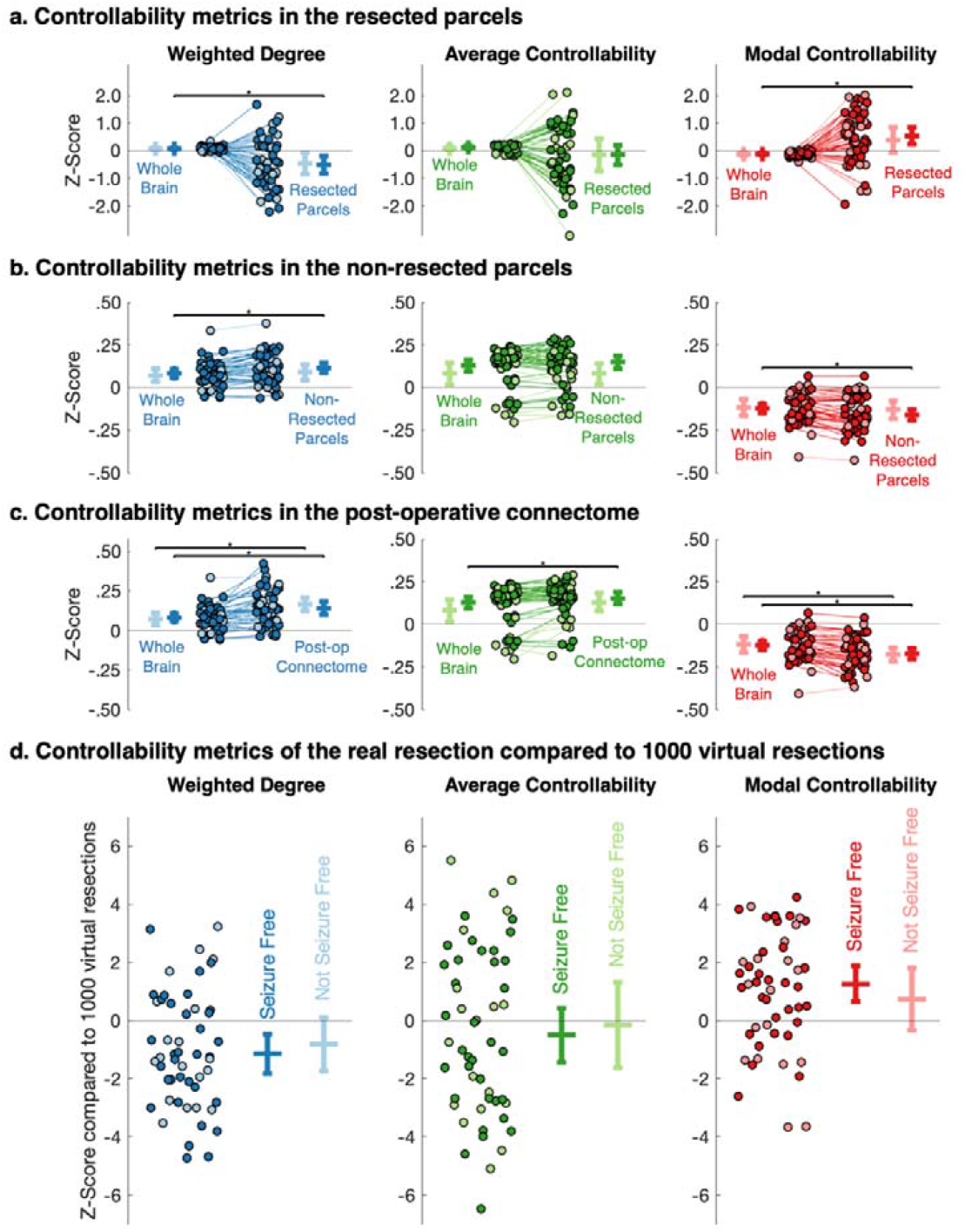
Resected parcels in seizure-free patients show lower weighted degree and higher modal controllability than equivalent parcels in controls and compared to virtual resections in patients. (a-c) Changes in the mean Z-scores for weighted degree, average controllability and modal controllability for each resective surgery patient between baseline, the (a) resected parcels, (b) the non-resected parcels and (c) the post-operative connectome. There were significant differences in the mean weighted degree (p = 0.002 and p= 6×10^−4^) and modal controllability (p = 6×10^−4^ and p= 4×10^−4^) for both resected and non-resected parcels in seizure free patients only (paired t-tests, adjusted for multiple comparisons using the Benjamini-Hochberg FDR method). For the post-operative connectome, there were significant differences in the mean weighted degree for both seizure free and not seizure free patients (p = 5×10^−4^ and p = 3×10^−5^, respectively), mean average controllability in seizure free patients only (p = 0.001) and mean modal controllability for both seizure free and not seizure free patients (p=3×10^−6^ and p=4×10^−5^, respectively) (paired t-tests, adjusted for multiple comparisons using the Benjamini-Hochberg FDR method). (d) Z scores of weighted degree, average controllability and modal controllability for the real resection compared to 1000 virtual resections for each of the patients, stratified by post-operative outcome. Bars show mean +/− 95% confidence intervals. The real resection parcels were significantly different from 0 for weighted degree (p=0.007) and modal controllability (p=0.002) in seizure free patients only (one sample t-tests, adjusted for multiple comparisons using the Benjamini-Hochberg FDR method).

The non-resected, remaining parcels had a higher mean weighted degree (p=6×10^−4^) and lower mean modal controllability (p=4×10^−4^), again only in patients that were seizure free following surgery (Figure 6b). To confirm this effect, we also generated the connectivity matrices again on the pre-operative scans, excluding the resection volumes from the possible streamlines (see Methods for details). We found that the post-operative connectome had a higher mean weighted degree and lower mean modal controllability but these findings were significantly different for patients that were both seizure free (p=5×10^−4^ and p=3×10^−6^) and not seizure free (p=3×10^−5^ and p=4×10^−5^) following correction for multiple comparisons. There was also an increase in average controllability in those that were seizure free (p=0.001) (Figure 6c).

## Discussion

### Drug-resistant epilepsy in children is associated with higher whole brain weighted degree, average controllability and modal controllability

In this diffusion MRI study of 79 children with drug-resistant epilepsy, we identified increases in network mean weighted degree, average and modal controllability in patients compared to controls (Figure 2). The changes in modal controllability have not previously been explored in the context of drug-resistant epilepsy. The graded increase between patients undergoing resective surgery (clinically characterised as unifocal epilepsy) and VNS implantation (clinically characterised as multifocal epilepsy) even when age and cognitive effects are corrected for, suggests that there are underlying network architectural differences in children with drug-resistant epilepsy that result in the brain being more easily able to reach difficult-to-reach states of neurophysiological activity including a seizure, which is defined as ‘abnormal excessive or synchronous neuronal activity in the brain’.^25^

### Drug-resistant epilepsy is associated with a loss of specialisation for dynamic roles

We found indicators of a loss of topographic specialisation in brain networks of patients with epilepsy in terms of the role of individual brain areas in shaping whole-brain dynamics. Wu-Yan *et al*. suggest that a strong correlation between average and modal controllability may represent a ‘specialisation’ in brain areas, which have either high modal or high average controllability (but not both)^17^. In our cohort of patients with drug-resistant epilepsy, this relationship was weaker, with increased (less negative) AC-MC correlation coefficients (Figure 3).

The mechanistic simulations we undertook allowed us to probe the nature of this disorganisation. By exploring differences in the structural connectivity matrices and network properties (Supplementary Figures S2 & S4), we hypothesized that these changes may occur due to increased connectivity in the epileptic brain, specifically in thalamocortical connectivity. Selectively increasing thalamocortical connections in control subjects by a low number of streamlines recapitulated many of the network properties seen in patients with epilepsy; including increases in average and modal controllability, weakening of the AC-MC correlation coefficient, decreased modularity and increased diffusion efficiency (Figure 4). This observation is in agreement with other studies that have found increased thalamocortical connectivity in children with drug-resistant epilepsy.^26^ Elucidating the nature of these changes may be important for complex system-based approaches to epilepsy treatment such that our interventions, at any layer of the complex system hierarchy, can reverse these changes back towards that of a ‘normal’ healthy network.^1,27^

A key question that arises from this set of findings is whether the increased thalamocortical connectivity and the resultant impacts on controllability are mechanisms for the epilepsy or are a result of uncontrolled seizures in this drug-resistant population. Epilepsy is also associated with a plethora of psychological and cognitive comorbidities, and it would be prudent to assess whether these and epileptic seizures are subserved by common underlying network alterations. Large longitudinal datasets from, for example, the ENIGMA consortium should be able to answer such questions.^28,29^

### Drug-resistant epilepsy is associated with decreased modal controllability in thalamic and posterior cingulate nodes

At the node-level, our study specifically identified decreased modal controllability in the cingulate cortex and thalamus of patients with drug-resistant epilepsy (Table 1). The cingulate cortex parcels have been shown to be part of the structural core of the cortex, with high node degrees and topographical centrality important, for functional integration.^30^ The thalamic parcels correspond to the locations of the centromedian and anterior nuclei, both established targets for deep brain stimulation as treatment for epileptic seizures; the anterior nucleus is thought to be a better target for focal epilepsy whereas the centromedian is thought to be a better target for generalised epilepsy (Supplementary Table 2).^31–33^ The involvement of multiple thalamic nuclei underscores the importance that thalamocortical circuitry plays in seizures and epilepsy.^34–37^ The increased connectivity of both regions lowers modal controllability values, perhaps indicating a reduced ability of these brain regions to move the brain into difficult-to-reach neurophysiological states (e.g., out of a seizure).

### Surgical resection is associated with parcels with higher modal controllability, specifically in seizure-free patients

The resected parcels in patients undergoing surgery and rendered seizure free had a lower mean weighted degree and higher mean modal controllability than the same parcels in controls (Figure 6a) and compared to virtual resection in patients (Figure 6d), suggesting an increased propensity for these parcels to reach difficult-to-reach neurophysiological states such as seizures. The finding of decreased weighted degree of these parcels complements previous studies that have noted variable connectivity patterns in focal cortical dysplasia, finding lesions with increased and decreased connectivity that corresponded to FCD type IIa and IIb, respectively.^38^ In our more heterogeneous cohort, we did not identify such differences in the weighted degree or controllability metrics between histological groups. Indeed, the direction of change of modal controllability seemed more consistent than the direction of change of weighted degree (Figure 6a), suggesting a propensity of these brain regions to move the brain into a seizure state.

### Limitations

Our study had several important technical limitations. Firstly, the cohorts were not age- or sex-matched, which may have been a particular issue given that the developmental trajectories of controllability metrics have not been fully elucidated.^23^ However, the patients had increased average and modal controllability despite being younger, thus if we extrapolate from previously described age effects, the effect size for group differences would have been diluted in our cohort. We used generalised linear model approaches to try adjust for some of these factors, including age, sex, and cognition. In these models, our findings of changes in controllability with age are consistent with Tang *et al*.’s findings that there is a stronger increase in the average controllability of whole brain networks with age compared to modal controllability.^23^ This strategy allowed us to maintain recording consistency in that all subjects were scanned on the same MRI scanner with identical protocol and post-processing, mitigating a number of otherwise confounding factors.

There were also limitations that have to do with interpretability of the findings. As recognised by previous studies, the use of network control theory to model and modify the human brain is still relatively new.^39–41^ Our findings that there are parcels with lower modal controllability in the thalamus and posterior cingulate regions and higher modal controllability in the resected regions are robust. The impact that these observations will have on both resective and neuromodulatory strategies for epilepsy is yet to be fully elucidated. It is foreseeable that this data could be used to inform neuromodulation targets as has recently been reported.^21,41^

### Conclusions

In this study, we have identified that paediatric drug-resistant epilepsy is associated with higher whole brain weighted degree, average controllability, and modal controllability and lower modal controllability of the thalamus and posterior cingulate parcels. In those undergoing resective surgery, modal controllability of the resected parcels is higher than would be expected and this effect is linked to post-operative outcomes. At the node level, children with drug-resistant epilepsy also have less strong correlations between weighted degree and controllability metrics, indicating a loss of topographic specialisation that may be partly due to increased thalamocortical connectivity. The findings emphasize the concept of more widespread changes being associated with ‘focal’ epilepsy and the importance of network-based approaches to better understand the pathophysiologic mechanisms and, in future, guide treatment. ^9,10^

## Methods

This retrospective study was conducted following approval from the Joint Research Office of Great Ormond Street Hospital & University College London Institute of Child Health (project ID 19BI26).

### Subjects

Subjects were eligible for inclusion if they had undergone volumetric T1MPRAGE and multi-shell diffusion imaging between 2015 and 2019 in one of three categories.

1. Resective Surgery: Patients who had undergone an MRI scan as part of the preoperative evaluation for resective epilepsy surgery. Surgery was carried out following discussion at a specialist epilepsy surgery multidisciplinary team meeting and some of these patients underwent intracranial evaluation with SEEG prior to proceeding with resective surgery. If there were multiple scans, the scan timepoint closest to the surgery (but prior to SEEG) was chosen. To reduce the impact of large structural abnormalities on the diffusion imaging metrics, patients were excluded if they had brain tumours, tuberous sclerosis, prior neurosurgical intervention or other large structural abnormalities such as previous strokes. Resections included a combination of anteromedial temporal lobe resections (27.0%) and extratemporal resections in the form of lesionectomies and lobectomies (73.0%), the exact extent of which was driven by the pre-surgical evaluation. Post-operative histology included focal cortical dysplasia and hippocampal sclerosis (Supplementary Table 1). Only those with a post-operative volumetric scan (to identify the resection volume) were included.
2. VNS: Patients who had undergone an MRI scan as part of the preoperative evaluation for a Vagal Nerve Stimulator. All patients were also discussed at a specialist epilepsy surgery multidisciplinary team meeting and deemed unsuitable for surgical resection due to presumed multifocal aetiology of seizures. To reduce the impact of large structural abnormalities on the diffusion imaging metrics, patients were excluded if they had tuberous sclerosis, prior neurosurgical intervention or other large structural abnormalities such as previous strokes.
3. Healthy Controls: Controls were scanned as part of another research study and were unaffected siblings of children with sickle cell disease. None of these children had epilepsy and the ones included had no significant MRI abnormalities on neuroradiological review.

Demographic information is available in Supplementary Table 1. Race and ethnicity in the groups was likely to be different but this data was not available for all subjects. However, race and ethnicity do not seem to have a major impact on grey and white matter volumes^42^ and, given the internal rankings used in the latter analyses (see below), this was not considered a major confound.

The entire workflow for image processing is summarised in Figure 7 and explained in detail below.

**Figure 7:**
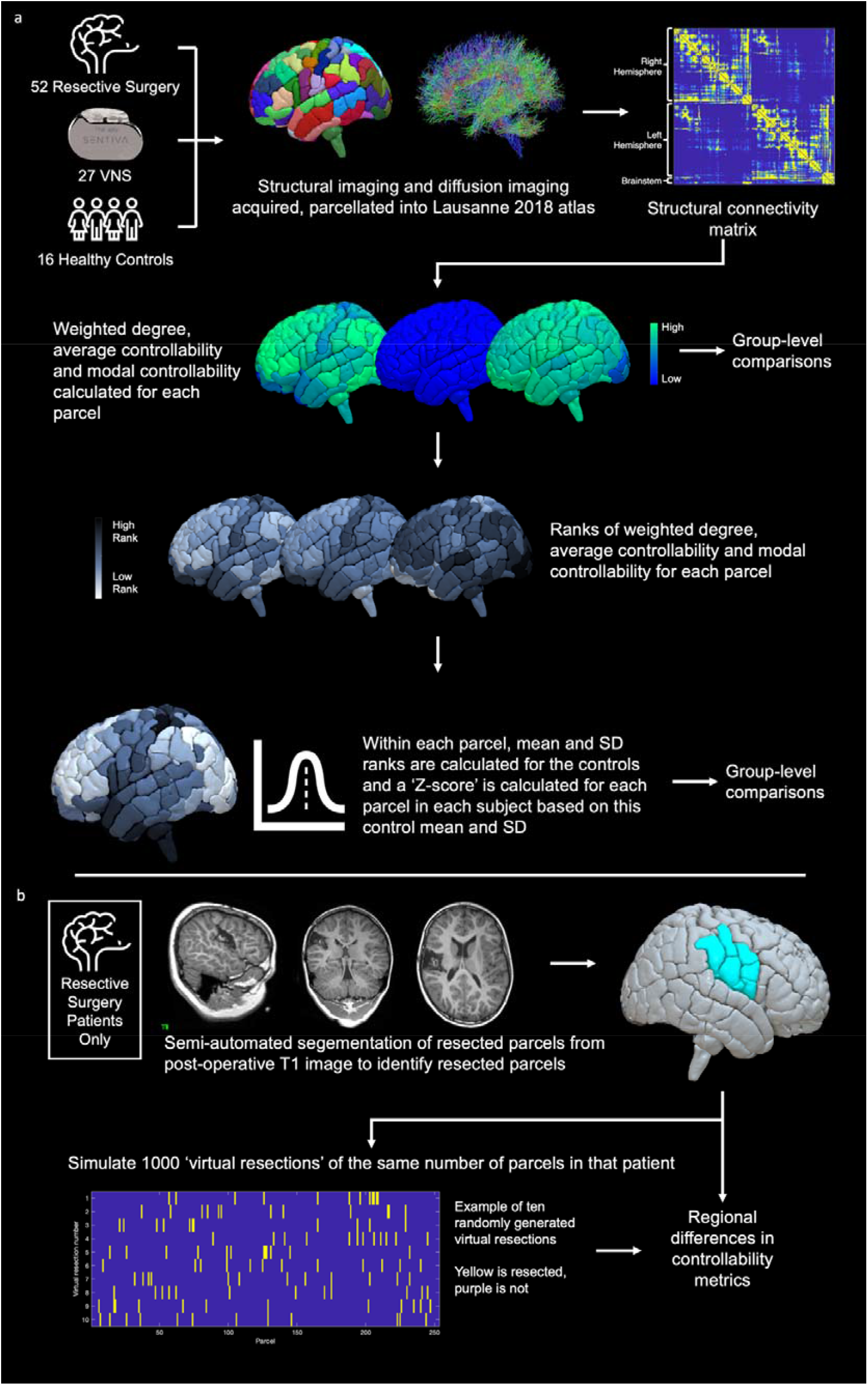
Summary of image processing workflow. (a) Following subject identification, structural imaging was parcellated into Scale 3 of the Lausanne 2018 resulting in 253 parcels per patient. Structural connectivity matrices were generated for each patient where the connection strength was derived from the number of streamlines between each of the 253 parcels. Using these matrices, the weighted degree, average controllability and modal controllability were calculated for each parcel, which were subsequently ranked within each subject and normalized based on the mean and standard deviation of the ranks in the healthy control subjects. (b) For the resective surgery cohort, resection volumes were mapped to the pre-operative parcellation to identify resected parcels. In order to robustly assess any regional differences, 1000 virtual resections of the same number of parcels as the actual resection were also mapped for each patient.

### MRI Scanning

All patients and controls were scanned on the same Siemens Magnetom Prisma 3.0T MRI scanner at Great Ormond Street Hospital, equipped with a 20-channel head coil. All scans (apart from the healthy controls) were acquired for routine clinical purposes.

The protocol included a T1 MPRAGE sequence and multi-shell diffusion sequence employing a diffusion-weighted spin-echo single shot 2D EPI acquisition, with multi-band radio frequency pulses to accelerate volume coverage along the slice direction. A multi-band factor of 2 was used to image 66 slices of 2 mm thickness with 0.2 mm slice gap. Diffusion gradients were applied over two ‘shells’: b = 1000 s/mm^2^ and b = 2200 s/mm^2^, with 60 non-collinear diffusion directions per shell in addition to 13 interleaved b = 0 (non-diffusion weighted) images. Other imaging parameters were: TR = 3050 ms, TE = 60 ms, field of view = 220 mm × 220 mm, matrix size = 110 × 110, in-plane voxel resolution = 2.0 mm × 2.0 mm, GRAPPA factor 2, phase-encoding (PE) partial Fourier = 6/8. An additional b = 0 scam was acquired, with identical readout to the diffusion-weighted scan, but with the phase encode direction flipped by 180° (in the anterior-posterior direction), for correction of susceptibility-related artifacts.

### MRI Processing

Structural parcellation was conducted using the T1 MPRAGE sequence processed using Connectome Mapper 3 ^43^, which is written in Python and uses Nipype.^44^ It is encapsulated in a BIDS app based in Docker and Singularity container technologies.^45–47^ Resampling to isotropic resolution, the Desikan-Killiany brain parcellation^48^ and brainstem parcellation^49^ were applied using FreeSurfer 6.0.1. Final parcellations were created by performing cortical brain parcellation at scale 3 of the Lausanne Atlas (v2018),^50^ probabilistic atlas-based segmentation of the thalamic nuclei^31^ and combination of all segmented structures, using in-house CMTK tools and the antsRegistrationSyNQuick tool of ANTS v2.2.0 ^51^, to create 253 parcels for each subject (219 cortical parcels; 30 subcortical structures including 7 thalamic nuclei on each side; 4 brainstem structures including midbrain, pons, medulla and superior cerebellar peduncle).

Connectomes were generated in mrtrix using in-house scripts. Following preprocessing (deionizing, bias correction), response was estimated using multi-shell, multi-tissue constrained spherical deconvolution. These diffusion images were registered to the structural images, optimized using SIFT2 and probabilistic tractography was performed using 5 million streamlines seeded from the entire white matter, resulting in symmetric, weighted, undirected connectivity matrices

All imaging was visually validated to verify the accuracy of the cortical segmentation and parcellation, ensure minimal movement artefact in the raw diffusion imaging and confirm accurate registration between the structural and diffusion images.

### Controllability Metrics

Connectivity matrices were analysed to ensure that the pre-requisites for controllability metrics were met, namely that no parcels or subnetworks were disconnected from the rest of the graph and that all diagonals were set to 0.^14^ No thresholding was applied, in line with SIFT2 recommendations.^52^ Average controllability and modal controllability were calculated for each parcel according to established methods.^14,15^ Briefly, we employed a simplified noise-free linear discrete-time and time-invariant network model to describe the dynamics of the brain:

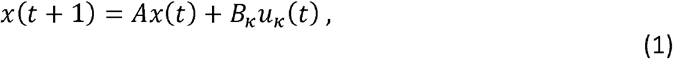

Where *x*: ℝ _≥ O_ → ℝ^*N*^ describes the state (i.e., activity level) of the parcels over time, and *A* ∈ ℝ^*N*×*N*^ is the structural connectivity matrix, which is subsequently stabilized by dividing by the mean edge weight. The input matrix *B*_*K*_ represents the control points *k*, where *k =* {*k*_1_, …, *k*_*m*_} and 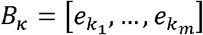, and *e*_*i*_ denotes the i-th canonical vector of dimension N. The input *u*_*k*_: ℝ _≥ O_ → ℝ^*m*^escribes the control strategy.

Average controllability of a network is equal to the average input energy from a set of control nodes and over all possible target states ^53,54^, which is proportional to the trace of the inverse of the controllability Gramian ----*Trace* 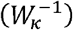, where:

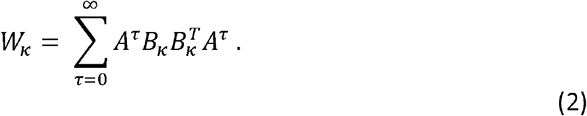

Regions with high average controllability tend to be densely connected, are involved in maintaining smooth and frequent transitions between related brain states and are enriched in the default mode network.^15^

Modal controllability refers to the ability of a node to control each evolutionary mode of a dynamic network.^55^ It is computed from the eigenvector matrix *V* = [*v*_*ij*_] of the structural connectivity matrix A. Specifically, modal controllability of the parcel *i* is defined as a scaled measure of the controllability of all N modes *λ*_1_ (*A*), …, *λ*_*N*_(*A*) from itself:

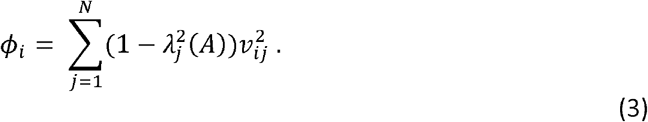

Regions of high modal controllability tend to be weakly connected areas, allow switching between brain states that have high energy barriers between them, and are enriched in frontoparietal and cingulo-opercular control systems.^15^

In addition, we calculated the weighted degree of each node, defined as the sum of weights of edges connected to the node.

### Graph Metrics

All graph metrics were calculated using the Brain Connectivity Toolbox.^56^ In order to probe the organisation of the graphs, metrics were chosen that were calculated on a whole graph basis and were assessed to reflect the level of organisation within the graphs.

- Density: Fraction of present connections to possible connections
- Modularity: The degree to which the network may be subdivided into clearly delineated groups according to the Louvain community detection algorithm.^57^
- Global Efficiency: Inverse of the mean shortest path length which is defined as the shortest distance between any pair of nodes. A measure of network efficiency that quantifies the ease of information transfer between 2 nodes; it requires knowledge of complete network structure.^58^
- Diffusion Efficiency: Inverse of mean first passage time which is defined as the expected number of steps it takes a random walker to reach one node from another. An alternative measure of network efficiency that quantifies the ease of information transfer between nodes but when this information spreads by random diffusion; it is based on limited local knowledge of network topology.^59^

### Statistical Analyses

At the network level, raw values of mean weighted degree, mean average controllability, mean modal controllability, the correlation coefficients and graph metrics were compared, correcting for age, sex, cognitive function (full scale IQ) and mean weighted degree using a generalized linear model (GLM) approach. The responses for mean modal controllability were not normally distributed (Shapiro-Wilk test) and so a gamma distribution was used for the GLM. Pairwise comparisons between the treatment groups were conducted using Fisher’s least significant difference procedure for the GLMs.

For the node-level analyses, ranks were normalized to the control mean and standard deviation to result in a Z-score for each parcel in each patient, which served to normalise the skewed distribution (Supplementary Figure S1). Further analyses were conducted on these Z-scores, comparing parcel-wise Z-scores across groups. Multiple comparisons were corrected for by choosing a Z-score threshold (+/− 3.1) that adequately corrected for the number of comparisons (253).

For the resective surgery group, post-operative scans were segmented using semi-automated segmentation on ITK-SNAP.^60^ Resection volumes were coregistered to the pre-operative scan parcellation to identify the parcels that had been resected (Figure 7). Any overlap (e.g. partially resected) was considered a resected parcel for the purposes of analysis. Between 3 and 24 parcels (median 12) were resected in each patient and there was an expected representation of resections from most parts of the brain, with the most common resections involving the anterior and mesial temporal structures and right frontal lobe with less resections involving the posterior mesial and occipital structures (Supplementary Figure S7). In addition, post-operative connectivity matrices were generated by re-running the tractography and excluding the resection volume using the mrtrix command tckexclude. The mean Z-scores of each metric (weighted degree, average and modal controllability) in the resected parcels, non-resected parcels and post-operative connectivity matrices were compared between groups using 2-tailed one or two sample t-tests, whilst correcting for multiple comparisons using the Benjamini-Hochberg FDR method.

### Simulation Modelling

Modelling was conducted using the structural connectivity matrices of the healthy controls as a base to attempt to transform them to a matrix with similar network properties to that of the patients with epilepsy. Prior analyses were used to inform the parameters of the changes applied to them.

In the first modelling exercise, the overall graph weight was increased by adding edges (where there were none before) that followed the same distribution as the edge weights for all subjects (Weibull Distribution with a = 0.717 and b = 0.325; using the curve fitting toolbox in Matlab, Supplementary Figure S3).

In the second modelling exercise, edge weights in the controls were increased by a low number of streamline counts, chosen at random from a distribution with mean of 50 and standard deviation of 15. This was informed by examining differences in the structural connectivity matrices between patients and controls (Supplementary Figure S5). The number of edges was fixed at 1440/31818, accounting for 4.5% of all edges in the graph, which was informed by the mean weighted degree & density increase between controls and patients. For each control, 10 anatomically constrained models were generated, where the weight was added only to randomly chosen thalamocortical edges. In addition, for each control, 200 anatomically unconstrained null models were generated, where weight was added to randomly chosen ipsilateral connections. Z-scores were generated for each controllability and graph metric within each subject from the distribution (mean and standard deviation) of the metrics in the null models and these were used to calculate the Z-scores for metrics in the anatomically constrained models and baseline connectomes. Z-scores were compared between groups using 2-tailed two sample t-tests and corrected for multiple comparisons using the Benjamini-Hochberg FDR method.

### Software, code, figures & data availability

All statistical analyses were performed on Matlab v 2018b and SPSS v24. All code and structural connectivity matrices are available on github (www.github.com/aswinchari/NetworkControl). Figures were made using Matlab and brain images were made using SurfIce, placed onto a parcellated paediatric average t1 brain (4.5-18.5y).^61^

### Citation Diversity Statement

Recent work in neuroscience has identified a bias in citation practices such that papers from women and other minority scholars are under-cited relative to the number of such papers in the field.^62,63^ Here we sought to proactively consider choosing references that reflect the diversity of the field in thought, form of contribution, gender, race, ethnicity, and other factors. First, we obtained the predicted gender of the first and last author of each reference by using databases that store the probability of a first name being carried by a woman. By this measure (and excluding self-citations to the first and last authors of our current paper), our references contain 10.63% woman(first)/woman(last), 21.61% man/woman, 13.45% woman/man, and 54.31% man/man. A comparison to leading neuroscience journals is show in Supplementary Figure S8. This method is limited in that a) names, pronouns, and social media profiles used to construct the databases may not, in every case, be indicative of gender identity and b) it cannot account for intersex, non-binary, or transgender people. Second, we obtained predicted racial/ethnic category of the first and last author of each reference by databases that store the probability of a first and last name being carried by an author of color. By this measure (and excluding self-citations), our references contain 11.22% author of color (first)/author of color(last), 19.93% white author/author of color, 23.53% author of color/white author, and 45.32% white author/white author. This method is limited in that a) names and Florida Voter Data to make the predictions may not be indicative of racial/ethnic identity, and b) it cannot account for Indigenous and mixed-race authors, or those who may face differential biases due to the ambiguous racialization or ethnicization of their names. We look forward to future work that could help us to better understand how to support equitable practices in science.

## Data Availability

All code and structural connectivity matrices are available on github (www.github.com/aswinchari/NetworkControl).

## Notes

**Funding:** AC is supported by a Great Ormond Street Hospital (GOSH) Children’s Charity Surgeon Scientist Fellowship. This work has been supported by the GOSH-National Institute of Health Research Biomedical Research Centre.

### Competing Interest Statement

The authors have declared no competing interest.

### Funding Statement

AC is supported by a Great Ormond Street Hospital (GOSH) Childrens Charity Surgeon Scientist Fellowship. This work has been supported by the GOSH-National Institute of Health Research Biomedical Research Centre.

### Author Declarations

This retrospective study was conducted following approval from the Joint Research Office of Great Ormond Street Hospital & University College London Institute of Child Health (project ID 19BI26). As it was a retrospective study using routinely collected clinical data, formal ethical approval was waived by the Joint Research Office of Great Ormond Street Hospital & University College London Institute of Child Health.

## References

1. Scott, R. C. Network science for the identification of novel therapeutic targets in epilepsy. F1000Research 5, 893 (2016).

2. Scott, R. C. et al. WONOEP APPRAISAL: The many facets of epilepsy networks. Epilepsia 59, 1475–1483 (2018).

3. Khambhati, A. N., Davis, K. A., Lucas, T. H., Litt, B. & Bassett, D. S. Virtual Cortical Resection Reveals Push-Pull Network Control Preceding Seizure Evolution. Neuron 91, 1170–1182 (2016).

4. Shah, P. et al. Characterizing the role of the structural connectome in seizure dynamics. Brain (2019) doi:10.1093/brain/awz125.

5. Kini, L. G. et al. Virtual resection predicts surgical outcome for drug-resistant epilepsy. Brain 142, 3892–3905 (2019).

6. Stacey, W. et al. Emerging roles of network analysis for epilepsy. Epilepsy Res. 159, 106255 (2020).

7. Pegg, E. J., Taylor, J. R., Keller, S. S. & Mohanraj, R. Interictal structural and functional connectivity in idiopathic generalized epilepsy: A systematic review of graph theoretical studies. Epilepsy & Behavior 106, 107013 (2020).

8. Meisel, C. Antiepileptic drugs induce subcritical dynamics in human cortical networks. 1904.13026 [q-bio] (2019).

9. Larivière, S. et al. Microstructure-Informed Connectomics: Enriching Large-Scale Descriptions of Healthy and Diseased Brains. Brain Connect 9, 113–127 (2019).

10. Larivière, S., Bernasconi, A., Bernasconi, N. & Bernhardt, B. C. Connectome biomarkers of drug-resistant epilepsy. Epilepsia (2020) doi:10.1111/epi.16753.

11. Besson, P. et al. Anatomic consistencies across epilepsies: a stereotactic-EEG informed high-resolution structural connectivity study. Brain 140, 2639–2652 (2017).

12. Proix, T., Jirsa, V. K., Bartolomei, F., Guye, M. & Truccolo, W. Predicting the spatiotemporal diversity of seizure propagation and termination in human focal epilepsy. Nature Communications 9, (2018).

13. Morgan, V. L., Rogers, B. P., González, H. F. J., Goodale, S. E. & Englot, D. J. Characterization of postsurgical functional connectivity changes in temporal lobe epilepsy. J Neurosurg 1–11 (2019) doi:10.3171/2019.3.JNS19350.

14. Karrer, T. M. et al. A practical guide to methodological considerations in the controllability of structural brain networks. J. Neural Eng. 17, 026031 (2020).

15. Gu, S. et al. Controllability of structural brain networks. Nature Communications 6, 8414 (2015).

16. Bassett, D. S., Khambhati, A. N. & Grafton, S. T. Emerging Frontiers of Neuroengineering: A Network Science of Brain Connectivity. Annual Review of Biomedical Engineering 19, 327–352 (2017).

17. Wu-Yan, E. et al. Benchmarking Measures of Network Controllability on Canonical Graph Models. J Nonlinear Sci 30, 2195–2233 (2020).

18. Haneef, Z. & Chiang, S. Clinical correlates of graph theory findings in temporal lobe epilepsy. Seizure - European Journal of Epilepsy 23, 809–818 (2014).

19. Bonilha, L. et al. Presurgical connectome and postsurgical seizure control in temporal lobe epilepsy. Neurology 81, 1704–1710 (2013).

20. Bernhardt, B. C. et al. Temporal lobe epilepsy: Hippocampal pathology modulates connectome topology and controllability. Neurology 92, e2209–e2220 (2019).

21. Scheid, B. H. et al. Time-evolving controllability of effective connectivity networks during seizure progression. PNAS 118, (2021).

22. Kwan, P. et al. Definition of drug resistant epilepsy: Consensus proposal by the ad hoc Task Force of the ILAE Commission on Therapeutic Strategies. Epilepsia 51, 1069–1077 (2010).

23. Tang, E. et al. Developmental increases in white matter network controllability support a growing diversity of brain dynamics. Nat Commun 8, 1–16 (2017).

24. Pedersen, M., Omidvarnia, A. H., Walz, J. M. & Jackson, G. D. Increased segregation of brain networks in focal epilepsy: An fMRI graph theory finding. Neuroimage Clin 8, 536– 542 (2015).

25. Fisher, R. S. et al. Epileptic Seizures and Epilepsy: Definitions Proposed by the International League Against Epilepsy (ILAE) and the International Bureau for Epilepsy (IBE). Epilepsia 46, 470–472 (2005).

26. Piper, R. J. et al. Functional connectivity of the anterior nucleus of the thalamus in pediatric focal epilepsy. Front. Neurol. 12, (2021).

27. Chari, A., Thornton, R. C., Tisdall, M. M. & Scott, R. C. Microelectrode recordings in human epilepsy: A case for clinical translation? Brain Commun (2020) doi:10.1093/braincomms/fcaa082.

28. Whelan, C. D. et al. Structural brain abnormalities in the common epilepsies assessed in a worldwide ENIGMA study. Brain 141, 391–408 (2018).

29. Larivière, S. et al. Network-based atrophy modeling in the common epilepsies: A worldwide ENIGMA study. Science Advances 6, eabc6457 (2020).

30. Hagmann, P. et al. Mapping the Structural Core of Human Cerebral Cortex. PLOS Biology 6, e159 (2008).

31. Najdenovska, E. et al. In-vivo probabilistic atlas of human thalamic nuclei based on diffusion-weighted magnetic resonance imaging. Scientific Data 5, 180270 (2018).

32. Li, M. C. H. & Cook, M. J. Deep brain stimulation for drug-resistant epilepsy. Epilepsia 59, 273–290 (2018).

33. Ilyas, A., Pizarro, D., Romeo, A. K., Riley, K. O. & Pati, S. The centromedian nucleus: Anatomy, physiology, and clinical implications. Journal of Clinical Neuroscience 63, 1–7 (2019).

34. Blumenfeld, H. The thalamus and seizures. Arch Neurol 59, 135–137 (2002).

35. He, X., Doucet, G. E., Sperling, M., Sharan, A. & Tracy, J. I. Reduced thalamocortical functional connectivity in temporal lobe epilepsy. Epilepsia 56, 1571–1579 (2015).

36. Kim, J. B. et al. Altered thalamocortical functional connectivity in idiopathic generalized epilepsy. Epilepsia 55, 592–600 (2014).

37. Peng, S.-J. & Hsin, Y.-L. Altered structural and functional thalamocortical networks in secondarily generalized extratemporal lobe seizures. Neuroimage Clin 13, 55–61 (2016).

38. Hong, S.-J. et al. A connectome-based mechanistic model of focal cortical dysplasia. Brain 142, 688–699 (2019).

39. Tu, C. et al. Warnings and caveats in brain controllability. NeuroImage 176, 83–91 (2018).

40. Pasqualetti, F., Gu, S. & Bassett, D. S. RE: Warnings and caveats in brain controllability. NeuroImage 197, 586–588 (2019).

41. Khambhati, A. N. et al. Functional control of electrophysiological network architecture using direct neurostimulation in humans. Netw Neurosci 3, 848–877 (2019).

42. Pfefferbaum, A. et al. Adolescent Development of Cortical and White Matter Structure in the NCANDA Sample: Role of Sex, Ethnicity, Puberty, and Alcohol Drinking. Cereb Cortex 26, 4101–4121 (2016).

43. Tourbier, S. et al. connectomicslab/connectomemapper3: Connectome Mapper v3.0.0-beta-RC1. (Zenodo, 2020). doi:10.5281/zenodo.3727708.

44. Gorgolewski, K. et al. Nipype: a flexible, lightweight and extensible neuroimaging data processing framework in python. Front Neuroinform 5, 13 (2011).

45. Gorgolewski, K. J. et al. BIDS apps: Improving ease of use, accessibility, and reproducibility of neuroimaging data analysis methods. PLOS Computational Biology 13, e1005209 (2017).

46. Docker: Lightweight Linux Containers for Consistent Development and Deployment | Linux Journal. https://www.linuxjournal.com/content/docker-lightweight-linux-containers-consistent-development-and-deployment.

47. Kurtzer, G. M., Sochat, V. & Bauer, M. W. Singularity: Scientific containers for mobility of compute. PLOS ONE 12, e0177459 (2017).

48. Desikan, R. S. et al. An automated labeling system for subdividing the human cerebral cortex on MRI scans into gyral based regions of interest. NeuroImage 31, 968–980 (2006).

49. Iglesias, J. E. et al. Bayesian segmentation of brainstem structures in MRI. Neuroimage 113, 184–195 (2015).

50. Cammoun, L. et al. Mapping the human connectome at multiple scales with diffusion spectrum MRI. J. Neurosci. Methods 203, 386–397 (2012).

51. Avants, B., Epstein, C. L., Grossman, M. & Gee, J. Symmetric diffeomorphic image registration with cross-correlation: evaluating automated labeling of elderly and neurodegenerative brain. Med Image Anal 12, 26–41 (2008).

52. Smith, R. E., Tournier, J.-D., Calamante, F. & Connelly, A. SIFT2: Enabling dense quantitative assessment of brain white matter connectivity using streamlines tractography. Neuroimage 119, 338–351 (2015).

53. Marx, B., Koenig, D. & Georges, D. Optimal Sensor and Actuator Location for Descriptor Systems using Generalized Gramians and Balanced Realizations. in vol. 3 2729–2734 vol.3 (2004).

54. Shaker, H. R. & Tahavori, M. Optimal sensor and actuator location for unstable systems. Journal of Vibration and Control 19, 1915–1920 (2013).

55. Hamdan, A. M. A. & Nayfeh, A. H. Measures of modal controllability and observability for first-and second-order linear systems. Journal of Guidance, Control, and Dynamics 12, 421–428 (1989).

56. Rubinov, M. & Sporns, O. Complex network measures of brain connectivity: uses and interpretations. Neuroimage 52, 1059–1069 (2010).

57. Blondel, V. D., Guillaume, J.-L., Lambiotte, R. & Lefebvre, E. Fast unfolding of communities in large networks. J. Stat. Mech. 2008, P10008 (2008).

58. Latora, V. & Marchiori, M. Efficient behavior of small-world networks. Phys Rev Lett 87, 198701 (2001).

59. Goñi, J. et al. Exploring the Morphospace of Communication Efficiency in Complex Networks. PLOS ONE 8, e58070 (2013).

60. Yushkevich, P. A. et al. User-guided 3D active contour segmentation of anatomical structures: significantly improved efficiency and reliability. Neuroimage 31, 1116–1128 (2006).

61. Fonov, V. et al. Unbiased average age-appropriate atlases for pediatric studies. Neuroimage 54, 313–327 (2011).

62. Bertolero, M. A. et al. Racial and ethnic imbalance in neuroscience reference lists and intersections with gender. bioRxiv 2020.10.12.336230 (2020) doi:10.1101/2020.10.12.336230.

63. Dworkin, J. D. et al. The extent and drivers of gender imbalance in neuroscience reference lists. Nat Neurosci 23, 918–926 (2020).

