## Supplementary Material for "Drug-resistant focal epilepsy in children is associated with increased modal controllability of the whole brain and epileptogenic regions"

**Supplementary Tables**

| Subject | Group | Sex | Cognitive Class | Outcome | Histology |
| --- | --- | --- | --- | --- | --- |
| 1 | Control | 'F' | 4 |  |  |
| 2 | Control | 'F' | 4 |  |  |
| 3 | Control | 'F' | 3 |  |  |
| 4 | Control | 'M' | 3 |  |  |
| 5 | Control | 'F' | 4 |  |  |
| 6 | Control | 'M' | 3 |  |  |
| 7 | Control | 'F' | 3 |  |  |
| 8 | Control | 'F' | 4 |  |  |
| 9 | Control | 'F' | 2 |  |  |
| 10 | Control | 'F' | 3 |  |  |
| 11 | Control | 'F' | 2 |  |  |
| 12 | Control | 'F' | 3 |  |  |
| 13 | Control | 'M' | 3 |  |  |
| 14 | Control | 'F' | 2 |  |  |
| 15 | Control | 'M' | 3 |  |  |
| 16 | Control | 'F' | N/A |  |  |
| 17 | Resection | 'M' | 4 | NSF | Non-diagnostic |
| 18 | Resection | 'F' | 1 | NSF | Hippocampal Sclerosis |
| 19 | Resection | 'M' | 1 | SF | Hippocampal Sclerosis |
| 20 | Resection | 'M' | 3 | SF | FCD Type 2b |
| 21 | Resection | 'M' | 1 | NSF | Hippocampal Sclerosis |
| 22 | Resection | 'M' | 1 | NSF | FCD Type 2a |
| 23 | Resection | 'M' | 1 | SF | FCD Type 2b |
| 24 | Resection | 'M' | 1 | NSF | Hippocampal Sclerosis |
| 25 | Resection | 'F' | 4 | SF | Non-diagnostic |
| 26 | Resection | 'M' | 4 | SF | Non-diagnostic |
| 27 | Resection | 'M' | 3 | SF | FCD Type 2b |
| 28 | Resection | 'M' | 1 | SF | Hippocampal Sclerosis |
| 29 | Resection | 'F' | 3 | NSF | Hippocampal Sclerosis |
| 30 | Resection | 'M' | 3 | SF | FCD Type 2b |
| 31 | Resection | 'M' | 2 | SF | FCD Type 2a |
| 32 | Resection | 'M' | 3 | SF | MCD |
| 33 | Resection | 'F' | 2 | SF | FCD Type 2b |
| 34 | Resection | 'F' | 1 | NSF | Hippocampal Sclerosis |
| 35 | Resection | 'M' | 1 | SF | FCD Type 2b |
| 36 | Resection | 'F' | 2 | SF | FCD Type 2b |
| 37 | Resection | 'F' | 2 | SF | Hippocampal Sclerosis |
| 38 | Resection | 'M' | 3 | SF | FCD Type 2b |
| 39 | Resection | 'M' | 3 | SF | Non-diagnostic |
| 40 | Resection | 'F' | N/A | NSF | Non-diagnostic |
| 41 | Resection | 'M' | 3 | SF | FCD Type 2b |
| 42 | Resection | 'M' | 1 | NSF | Non-diagnostic |
| 43 | Resection | 'F' | 1 | SF | FCD Type 2a |
| 44 | Resection | 'M' | 3 | NSF | FCD Type 2b |
| 45 | Resection | 'F' | 2 | SF | FCD Type 2b |
| 46 | Resection | 'F' | 4 | SF | Hippocampal Sclerosis |
| 47 | Resection | 'M' | 1 | NSF | FCD Type 2b |
| 48 | Resection | 'M' | 4 | SF | FCD Type 2b |
| 49 | Resection | 'M' | 2 | NSF | FCD Type 2b |
| 50 | Resection | 'F' | 1 | NSF | Non-diagnostic |
| 51 | Resection | 'M' | 3 | SF | Non-diagnostic |
| 52 | Resection | 'M' | 3 | NSF | Non-diagnostic |
| 53 | Resection | 'F' | 3 | SF | FCD Type 2b |
| 54 | Resection | 'F' | 2 | SF | FCD Type 2b |
| 55 | Resection | 'F' | 2 | SF | FCD Type 2b |
| 56 | Resection | 'F' | 1 | NSF | FCD Type 2b |
| 57 | Resection | 'M' | 3 | SF | Non-diagnostic |
| 58 | Resection | 'F' | 1 | SF | Non-diagnostic |
| 59 | Resection | 'M' | 3 | NSF | Non-diagnostic |
| 60 | Resection | 'M' | 1 | SF | Non-diagnostic |
| 61 | Resection | 'M' | 4 | SF | FCD Type 2b |
| 62 | Resection | 'F' | 4 | SF | FCD Type 2b |
| 63 | Resection | 'F' | 4 | NSF | Non-diagnostic |
| 64 | Resection | 'M' | 3 | SF | FCD Type 2a |
| 65 | Resection | 'F' | 3 | SF | FCD Type 2a |
| 66 | Resection | 'M' | 2 | NSF | Hippocampal Sclerosis |
| 67 | Resection | 'M' | 1 | SF | Hippocampal Sclerosis |
| 68 | Resection | 'M' | N/A | NSF | FCD Type 2b |
| 69 | VNS | 'M' | 1 | No Response | Non-diagnostic |
| 70 | VNS | 'F' | 1 | Response |  |
| 71 | VNS | 'M' | 4 | Response |  |
| 72 | VNS | 'M' | 1 | No Response |  |
| 73 | VNS | 'F' | 1 | No Response |  |
| 74 | VNS | 'M' | 3 | No Response |  |
| 75 | VNS | 'M' | 1 | Response |  |
| 76 | VNS | 'M' | 1 | Response |  |
| 77 | VNS | 'F' | 1 | No Response |  |
| 78 | VNS | 'M' | 3 | No Response |  |
| 79 | VNS | 'M' | N/A | No Response |  |
| 80 | VNS | 'M' | N/A | Response |  |
| 81 | VNS | 'M' | N/A | No Response |  |
| 82 | VNS | 'M' | N/A | Response |  |
| 83 | VNS | 'F' | 3 | Response |  |
| 84 | VNS | 'M' | 1 | Response |  |
| 85 | VNS | 'F' | 2 | No Response |  |
| 86 | VNS | 'F' | 1 | No Response |  |
| 87 | VNS | 'M' | 1 | Response |  |
| 88 | VNS | 'F' | 3 | No Response |  |
| 89 | VNS | 'M' | 3 | No Response |  |
| 90 | VNS | 'M' | 1 | No Response |  |
| 91 | VNS | 'M' | 1 | Response |  |
| 92 | VNS | 'M' | N/A | No Response |  |
| 93 | VNS | 'F' | 1 | Response |  |
| 94 | VNS | 'M' | 1 | No Response |  |
| 95 | VNS | 'M' | N/A | No Response |  |

*Supplementary Table 1: Group and demographic details of each participant. Cognitive class categories are 1 = >2SD below the mean, 2 = 1-2SD below the mean, 3 = 0-1 SD below the mean, 4 = 0-1 SD above the mean, N/A = not assessed. For VNS, response = >50% reduction in seizures. SF = seizure free, NSF = not seizure free, MCD = malformation of cortical development, FCD = focal cortical dysplasia*

| **Atlas Parcel** | **Terminologia Anatomica Nuclei** | **Function** |
| --- | --- | --- |
| anterior | Anterior | Output to cingulate/parahippocampal gyri; common target for DBS for epilepsy |
| central lateral, lateral posterior, medial pulvinar | CL & CM | Output to motor cortex and striatum; common target for DBS for epilepsy |
| ventral latero-ventral | VL & VPL | Motor (VL) and sensory/insula (VPL) projections; common targets for DBS for dystonia (VL) and pain (VPL) |
| ventral latero-ventral | LP |  |
| medio-dorsal | MD | Output to prefrontal cortex; targets for DBS for obsessive-compulsive disorder |

*Supplementary Table 2: Atlas-based thalamic nuclei which have increased modal controllability, their corresponding terminologia anatomica nuclei and function. CL = central latera, CM = centromedian, VL = ventral lateral, VPL = ventral posterolateral, LP = lateral posterior, MD = medial dorsal.*

**Supplementary Figures**

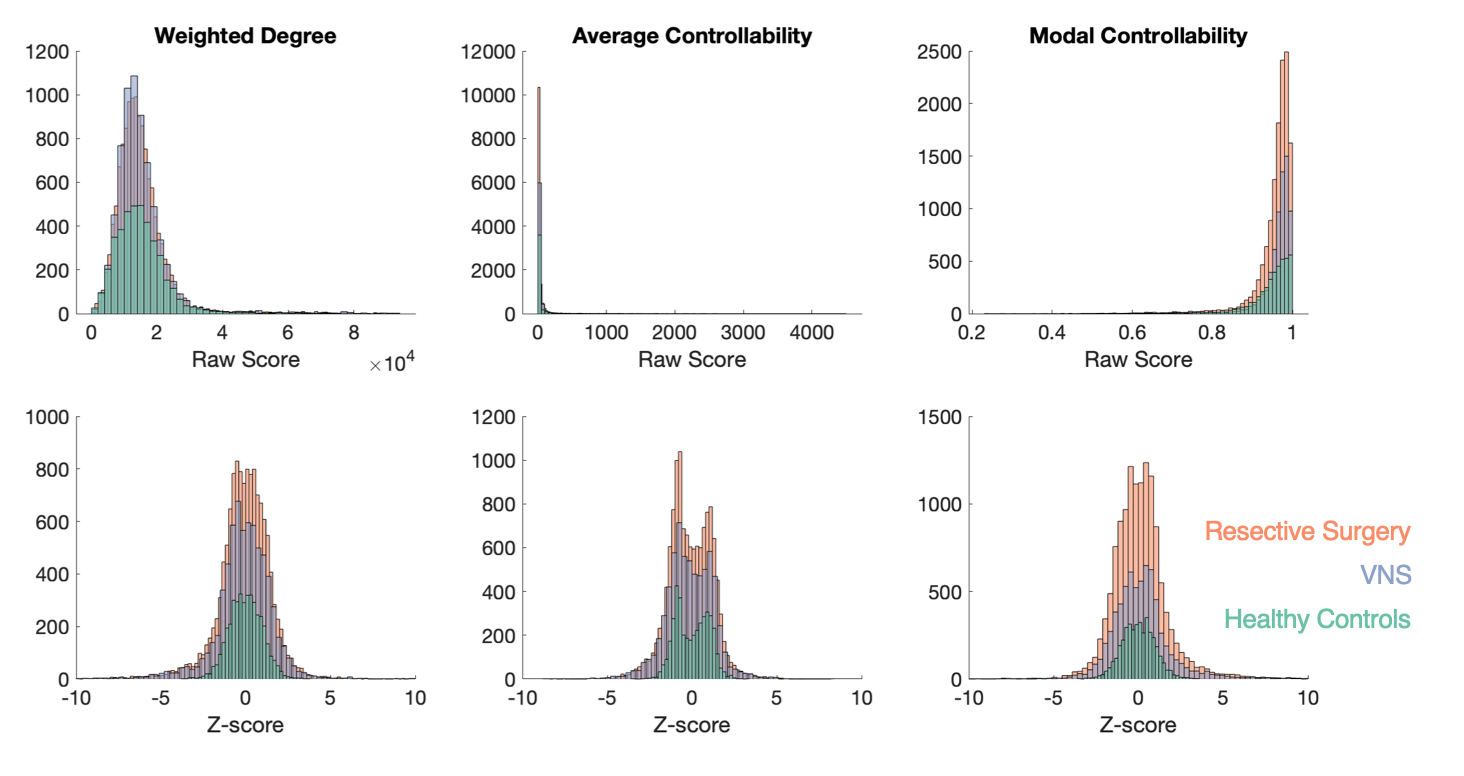

*Supplementary Figure S1: Histograms illustrating the non-parametric nature of the raw scores (top panel) which were normalised by converting each value to a rank within each individual patient and using the mean and standard deviation of the ranks in the healthy controls to calculate a Z-score for each parcel (bottom panel).*

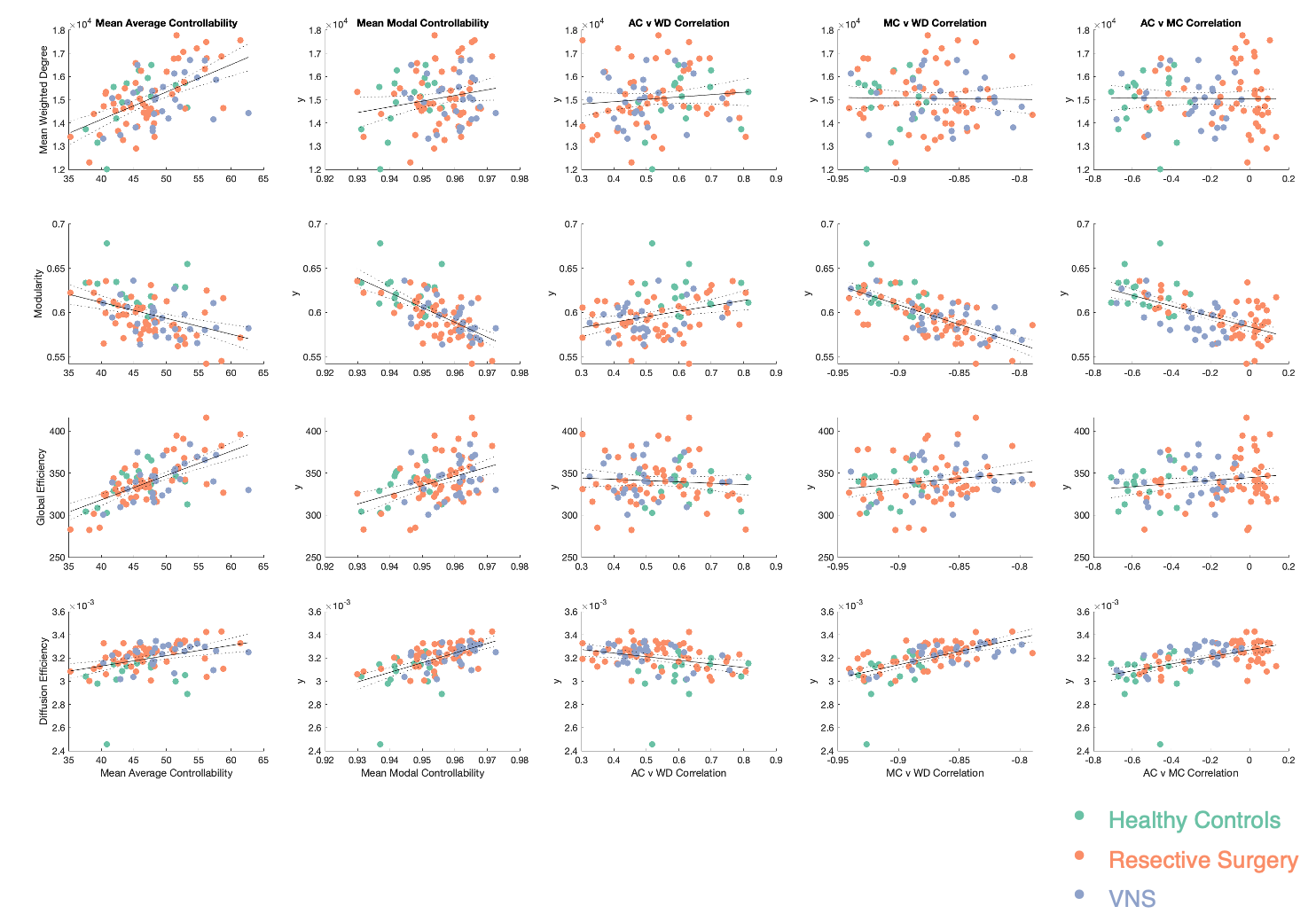

*Supplementary Figure S2: Relationships between five controllability variables and four traditional graph theory metrics for each of the patients. AC = average controllability, MC = modal controllability and WD = weighted degree. Lines show correlation coefficients between measures +/- 95% confidence intervals. Note that the groups seem to segregate best by AC vs MC correlation and none of the graph metrics correlate strongly with this measure.*

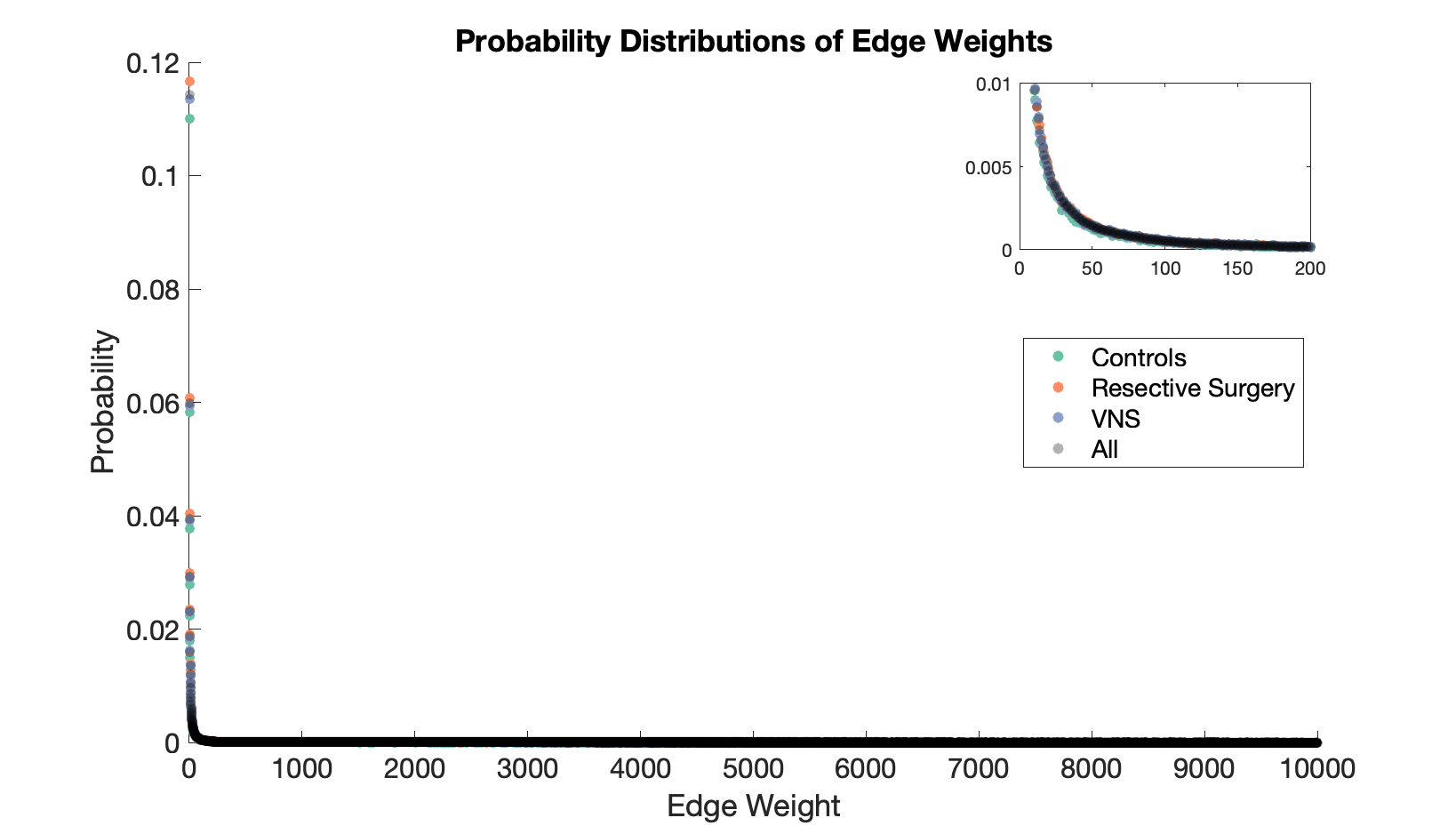

*Supplementary Figure S3: Probability distributions of the edge weights from the 3 groups of patients and all groups combined (in grey), showing similar distributions across the groups. Inset shows same graph zoomed in showing strong overlap between groups. The overall distribution curve was best fit using Weibull distribution with a = 0.717 and b = 0.325 (RMS of error 2.3 x 10^-5^), providing a better fit than an exponential distribution (RMS of error 3.4 x 10^-4^). Curves were fit using the curve fitting toolbox in matlab.*

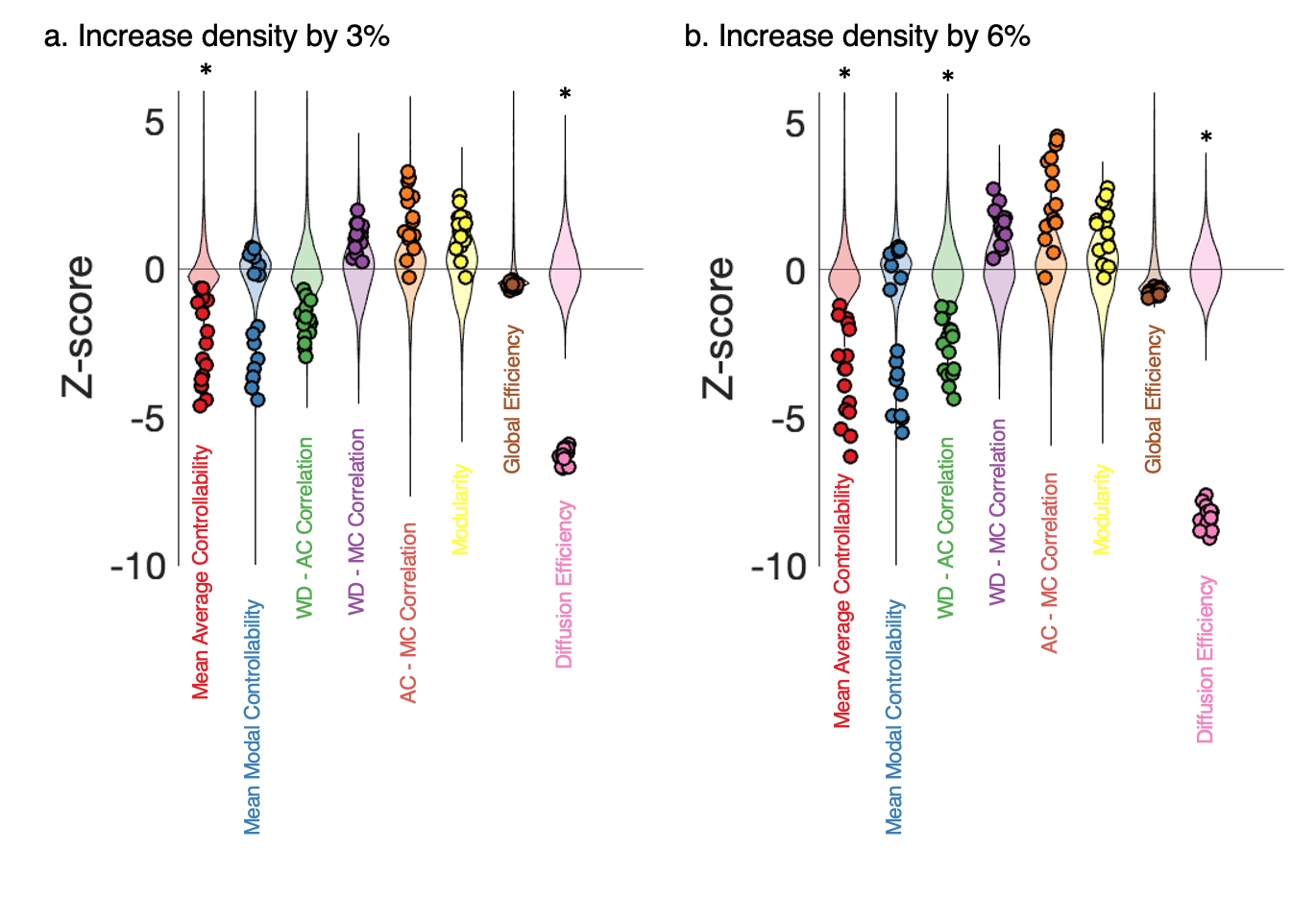

***Supplementary Figure S4: Increasing the density of connections in healthy controls at random does not explain the changes observed in children with drug-resistant epileps****. Z-scores of graph theory and controllability metrics of the original connectomes from the healthy controls compared to 3200 simulations (200 for each subject, shown as background violin plots) for each subject where extra edges were added. Adding edges (3% = 1000 edges, 6% = 2000 edges) leads to a statistically significant increase in average controllability and diffusion efficiency, and a trend towards increase in modal controllability and decrease in modularity. This does not recapitulate the changes seen in the patients with epilepsy. The WD-AC correlation significantly increases, WC-MC and AC-MC correlations do not change significantly, indicating that the ‘disorga. * indicates that average Z-score of the controls (n=16) compared to Z-scores from all the simulations (n=3200) is > 2.5 (effectively correcting for 8 multiple comparisons).*

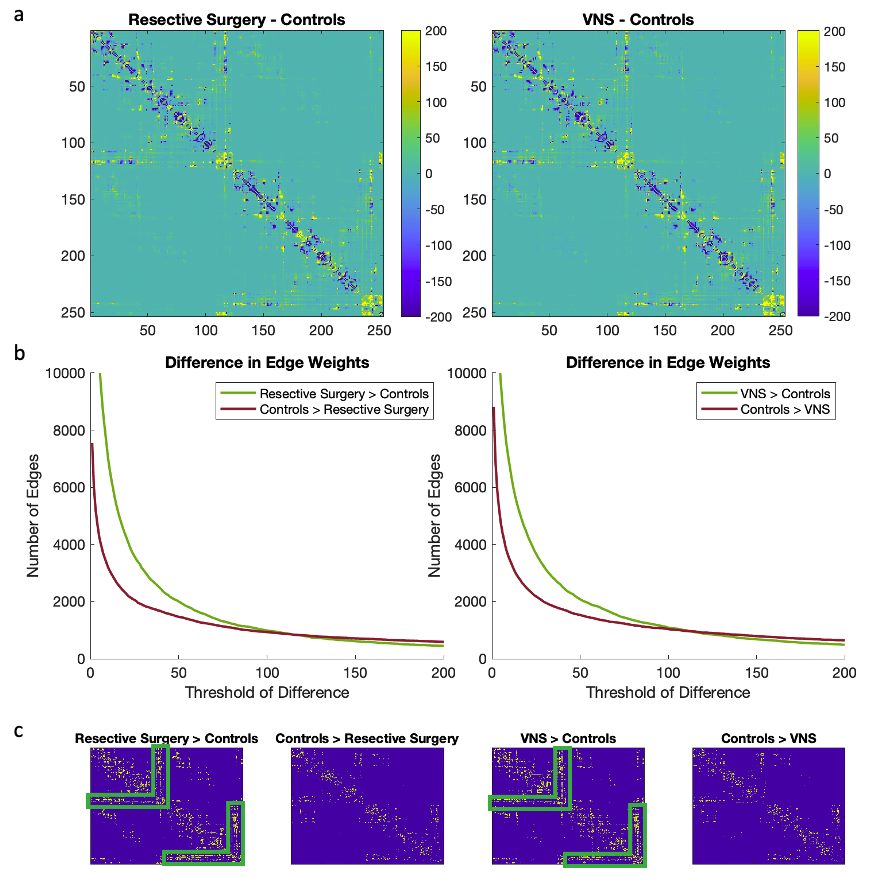

***Supplementary Figure S5: Average structural connectivity matrix differences between controls and patients with drug-resistant epilepsy.*** *(a) Average structural connectivity matrices for each group were constructed and the differences between resective surgery and controls (left) and VNS and controls (right) are shown. (b) Quantification of the number of edges different between controls and patients above a certain threshold. There were more low edge weight differences (>100 streamlines) in the patients and high weight edge differences (>100 streamlines) in the controls. (c) Location of these low edge weight difference connections (20-80 streamlines) in the structural connectivity matrices reveals that additional connections in patients were predominantly in ipsilateral thalamocortical connections (green boxes) with other scattered ipsilateral cortico-cortical connections having both increased and decreased weight in the patients compared to controls.*

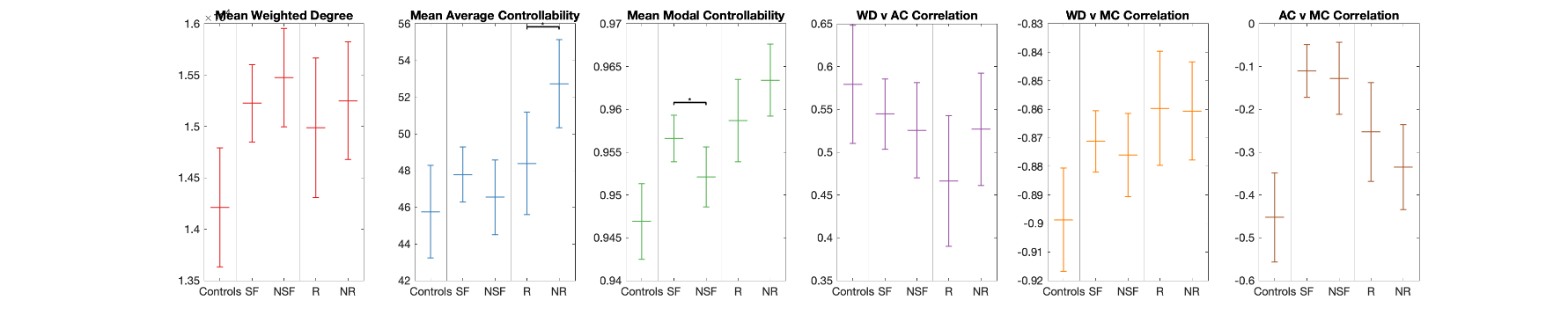

***Supplementary Figure S6: Global controllability do not associate with treatment outcome.*** *Estimated marginal means (95% Wald confidence interval) for the different groups, stratified by treatment outcomes. Following adjustment for age, sex, cognitive function and mean weighted degree, pairwise comparisons (adjusted for multiple comparisons) between groups resulted in there being small differences between the VNS responders and non-responders for mean average controllability (p=0.02) and seizure-free and non seizure-free resective surgery patients for mean modal controllability (p=0.05).*

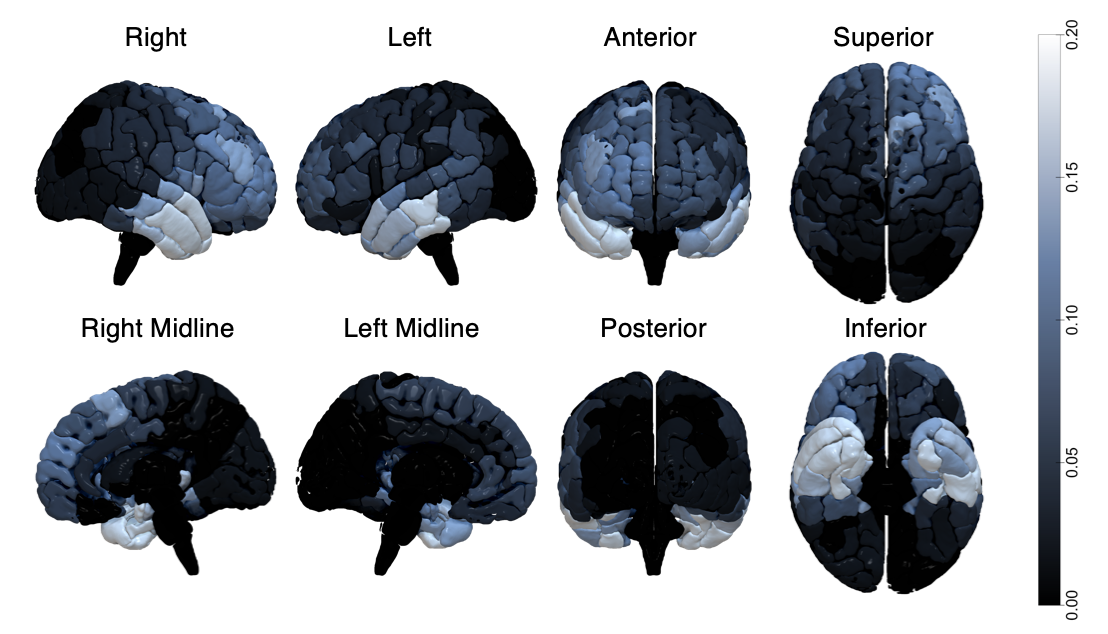

***Supplementary Figure S7: Composite figure of resected parcels across 52 patients.*** *Views of a template paediatric brain showing the proportion of parcels that were resected across the 52 resective surgery patients. As would be expected, the most commonly resected parcels were the anterior and mesial temporal lobe structures (right > left) with fewer resections involving midline and posterior parcels.*

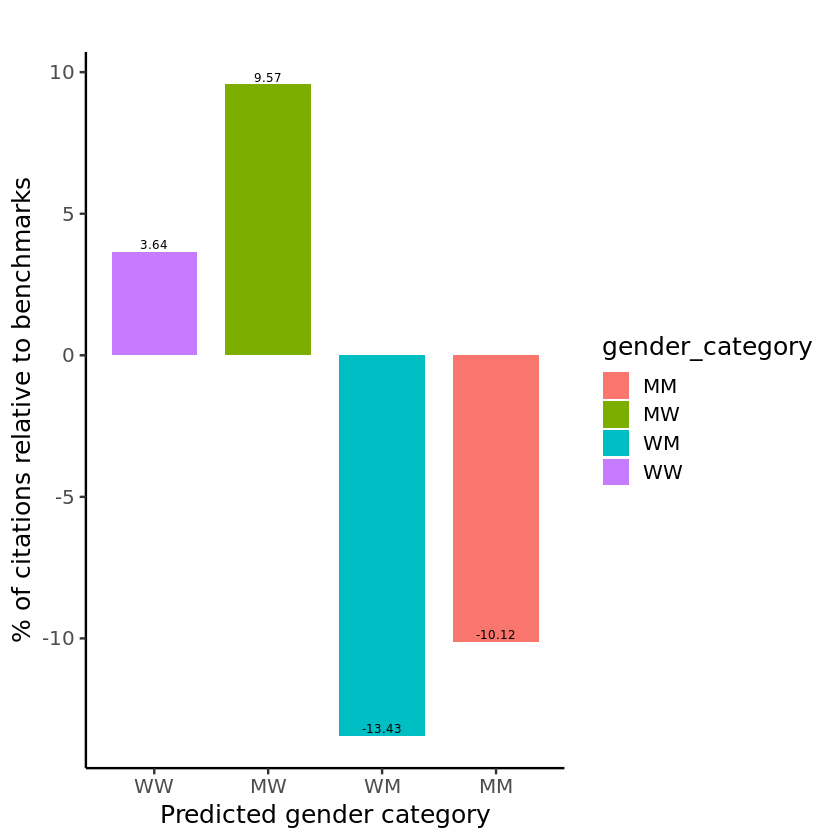

***Supplementary Figure S8: Citation diversity.*** *Percentage of citations above/below threshold from leading neuroscience journals of different predicted gender categories (M = man and W = woman)*
